# Mapping brain connections to inflammatory networks in depression identifies sex-specific neuroimmune networks and neurobiological subtypes

**DOI:** 10.64898/2026.08.01.26359430

**Authors:** Truc D. X. Chu, Lucy M. Hui, Nick Teller, Nasreen Khatri, Georgia E. Hodes, J. Jean Chen

**Author notes:** Co-first authors (Truc D. X. Chu, Lucy M. Hui). Co-senior authors (Georgia E. Hodes, J. Jean Chen).

## Abstract

**Background:** Major depressive disorder is a heterogeneous psychiatric condition, complicating clinical diagnosis and treatment. Efforts to stratify MDD have led to the utilization of systemic inflammatory and neuroimaging markers. Understanding of the relationship between these markers and how they correspond to depressive symptoms is crucial for identifying personalized metrics to rationally diagnose MDD.

**Methods:** The CANBIND1 dataset comprising cohorts of people with MDD (n=211) and age-matched healthy controls (n=122) was used. Weighted gene co-expression network analysis (WGCNA) was applied to cluster multiplex ELISA cytokine data and identify inflammatory modules. Microstructural metrics were derived from diffusion-weighted imaging (DWI) in the same individuals. The relationships among inflammatory modules, imaging markers, and clinical features were examined using correlation analysis.

**Results:** We demonstrate that networks of peripheral inflammatory markers relate to specific depressive symptoms. Additionally, these cytokine networks are associated with diffusion MRI metrics of tissue microstructure, especially the correlated diffusion index (CDI). Distinct patterns were observed in patients compared to age-matched controls. Notably, these associations are more pronounced in gray matter than white matter, and more in females than in males.

**Conclusion:** Our findings reveal sex-dependent networks within systemic inflammatory markers, which are in turn linked to sex-specific disease subtypes. This work provides a neurobiological framework which may facilitate the identification of biologically meaningful subtypes of depression, ultimately improving diagnosis and treatment.

## Introduction

While the brain can impact inflammation in the body, peripheral inflammation can also trigger neuroinflammation through various mechanisms. These include disruption of the blood-brain barrier (BBB), activation of glial cells, and effects on autonomic nerves (1). Systemic inflammatory stimuli can activate sensitized “primed” microglia in demyelinating diseases, leading to stronger and quicker pathological responses (2). The transmission or suppression of inflammation from the periphery to the central nervous system involves key components such as monocytes and microglia, involving signaling such as via the CD200-CD200R1 pathway.

Microglial activation plays a crucial role in neuroinflammation and is associated with various neurological disorders, including depression (3) and suicidal ideation (4,5). This activation can be triggered by systemic and central nervous system signals, leading to the release of cytokines and chemokines (3). Key cytokines and chemokines related to microglial activation include monocyte chemoattractant protein 1 (MCP-1/CCL-2), granulocyte macrophage colony stimulating factor (GM-CSF), and interleukin-1 (IL-1), which promote classical pro-inflammatory signaling by microglia, while IL-4, IL-10, and macrophage derived chemokine (MDC/CCL22) are associated with alternative anti-inflammatory microglia signaling (6). Activated microglia can disrupt the BBB and significantly increase the production and peripheral access of various cytokines and chemokines, particularly through interactions with astrocytes (7). This increases the ability of these inflammatory proteins to travel into the brain (8–10). Neuroinflammation could also precipitate BBB leakage and aggravate pathology through vascular dysfunction (11). Neuroinflammation can contribute to neuronal damage, demyelination, and the overall progression of neurodegeneration in autoimmune diseases(12,13).

Diffusion MRI (dMRI), which measures the rate and direction of water molecule movement in the brain, provides a means to determine changes in grey matter microstructure, trace white matter tracts, and identify regions of connectivity. By examining measures including mean diffusivity (MD) and fractional anisotropy (FA), we can identify regions in which microstructure has been impacted. Specifically, FA is related to degree of diffusion anisotropy, indicating how strongly water movement is constrained to a direction such as along nerve fibers. MD indicates the overall magnitude of diffusion. Reduced FA and increased MD often reflect a loss of tract integrity due to demyelination or tissue breakdown (14). Recent studies have also shown significant correlations between inflammation and diffusion MRI parameters across various physiological conditions. In non-demented elderly individuals, higher systemic inflammation was associated with lower FA and higher radial diffusivity in multiple white matter regions (15). Similarly, in virally suppressed HIV-infected individuals, elevated inflammatory biomarkers were associated with white matter abnormalities. Specifically, higher MCP-1 was associated with lower FA and higher MD across various tracts (16). More elaborate methods such as diffusion basis spectrum imaging (DBSI) was also used to demonstrate inflammation in multiple sclerosis (MS) lesions (17,18). Similarly, neuro-inflammation imaging was employed to assess white matter inflammation and damage in Alzheimer’s disease, revealing elevated cellular diffusivity in preclinical and early symptomatic phases (19). These data support that diffusion MRI metrics can reflect underlying inflammatory processes in different pathological conditions.

Although previous studies have suggested that diffusion MRI can detect microstructural changes associated with neuroinflammation (20–22), the relationship between peripheral inflammatory networks, regional brain microstructure, and specific depressive symptoms remains poorly understood. Here we analyzed multimodal data from the Canadian Biomarker Integration Network in Depression (CANBIND) (23), integrating peripheral inflammatory markers, diffusion MRI measures of brain microstructure, and clinical assessments. We took an agnostic data driven approach to determine whether distinct networks of systemic inflammatory cytokines are associated with specific depressive symptoms and whether these same cytokine networks correspond to regional differences in brain microstructure. By defining these brain–immune relationships, we try to clarify how inflammation contributes to different symptom dimensions and help identify biologically meaningful subtypes of MDD.

## Methods

### Data set

The study cohort was taken from the Canadian Biomarker Integration Network in Depression (CANBIND-1) (23). The cohort included individuals with major depressive disorder (MDD) and controls, whose distributions and data types were summarized in **Fig. 1** and **Table S1**. MDD diagnoses were established according to DSM-IV-TR criteria using a structured clinical interview (MINI) and rigorous inclusion and exclusion criteria defined by the CANBIND study protocol (24). CANBIND-1 was established to identify the interrelationship across the molecular, clinical and imaging markers. The molecular markers included in the current analysis consist of blood protein levels of cytokine and chemokines. The CANBIND-1 data was collected at 6 sites.

**Figure 1.**
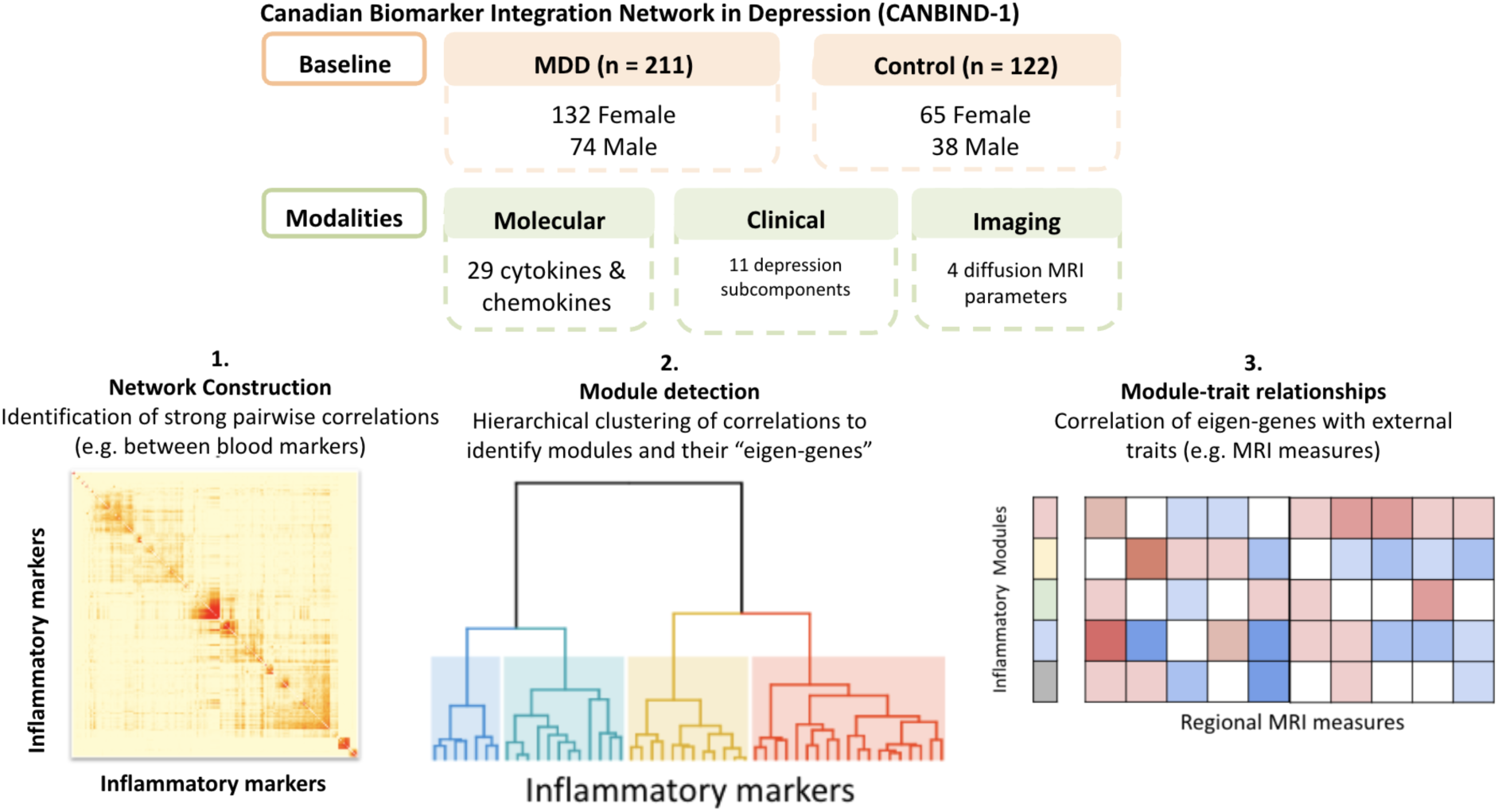
Data used in this study came from the CANBIND data which collected molecular, clinical and imaging data from 211 individuals with depression and 122 aged matched healthy controls (top). Data was analyzed using the WCGNA molecular data-analysis workflow to identify cytokine clusters. The eigen-gene value for each cluster was correlated to generate module to trait relationships with imaging and clinical data (bottom). Data was then clustered based on trait relationships to identify subtypes of depression in men and women.

### Inflammatory modules from molecular data

We performed weighted gene correlation network analysis (WGCNA) on the molecular markers (**Table S2.**) to better understand interactions among markers. WGCNA is a powerful method for analyzing gene co-expression networks and identifying functional modules, which is also applied to proteomic and metabolomic data (25,26). Inflammatory modules were extracted from the blood markers using WGCNA, separately for MDD and controls to help understand group-specific mechanisms (see Supple. Mat. for details). The Adjusted Rand Index (ARI) and Normalized Mirkin Metric (NMM) were used to quantify similarity and differences in cytokine module organization between networks.

### Clinical markers

The primary clinical marker for depression symptoms was the Montgomery-Åsberg Depression Rating Scale (MADRS). We also assessed the Dimensional Anhedonia Rating Scale (DARS) for anhedonia and the Pittsburgh Sleep Quality Index (PSQI) for sleep quality (see Supple. Mat. for details of each score). Scores were obtained for both the MDD and control groups.

### Imaging markers

The imaging protocol is described in Lam et al., 2016 (24). The imaging markers were derived from single-shell diffusion-weighted MRI data. These markers are mean diffusivity (MD), fractional anisotropy (FA), free-water corrected FA (FAt), free-water signal fraction (F), and correlated diffusion index (CDI) (see full details in Supple. Mat.).

### Data integration

The relationships between inflammatory modules and clinical scores were determined by correlating the module eigengenes with clinical markers to reveal potentially key biological processes for the modules, in this case using Spearman’s correlation.

Partial least squares structural equation modelling (PLS-SEM) (27) was used to investigate the mediation effects of peripheral inflammatory markers on the relationship between dMRI metrics and clinical symptoms. Furthermore, to identify data-driven subtypes within MDD groups, separated by sex, Similarity Network Fusion (SNF) (28) was performed (see full details in Supple. Mat.).

The imaging markers were submitted to multi-site harmonization using the ComBAT method (29). The WGCNA-derived inflammatory modules were correlated with the dMRI metrics in grey matter and white matter parcellations, obtained using FreeSurfer and the JHU atlas, respectively. The statistically significant correlations, corrected for FDR, were mapped to both the cortical and subcortical parcellations using R.

## Results

Analysis of circulating inflammatory markers indicated that that interleukin 1 receptor antagonist (IL1ra) (t = 3.5, FDR-adjusted p < 0.05) and monocyte chemoattractant protein 1 (MCP1/CCL2) (t = 3.21, FDR-adjusted p < 0.05) were the only individual inflammatory markers significantly elevated in the MDD group compared to controls (**Fig. S1**). No inflammatory markers were significantly lower in the MDD group than in the control group. Control and MDD scores follow significantly different distributions in their MADRS scores (**Fig.S2**). At the whole brain level there were no significant differences in dMRI metrics between MDD and control groups (**Fig. S3**).

Cytokines and chemokines work in concert in the body and brain to exert their effects. Therefore, we took an agnostic clustering approach commonly used on sequencing data, to further identify cooperative groups and patterns of inflammatory proteins (**Fig. 2**, **Table S3-S5**). The initial WGNA was performed across both males and females with MDD (male-female consensus). Each module was assigned an arbitrary color, and cytokines not assigned to a co-expression network are listed as the grey module by network analysis. ARI and NMM were then used to calculate cytokine co-expression similarity and differences regardless of module label by considering all pairwise relationships between cytokines (Table S6). Several inflammatory modules were moderately preserved across MDD and control participants, including modules centered on VEGF, IFN-γ, and IL-6 or IL-4 and eotaxin (ARI= 0.175/NMM= 0.239). However, the composition of these modules was moderately reorganized in MDD through the recruitment of additional growth factors (FGF and PDGF), immune signaling molecules involved in leukocyte activation and trafficking (GM-CSF, IL-15, and RANTES), and immunoregulatory cytokines (IL-10 and IL-5), indicating altered coordination of inflammatory signaling.

**Figure 2.**
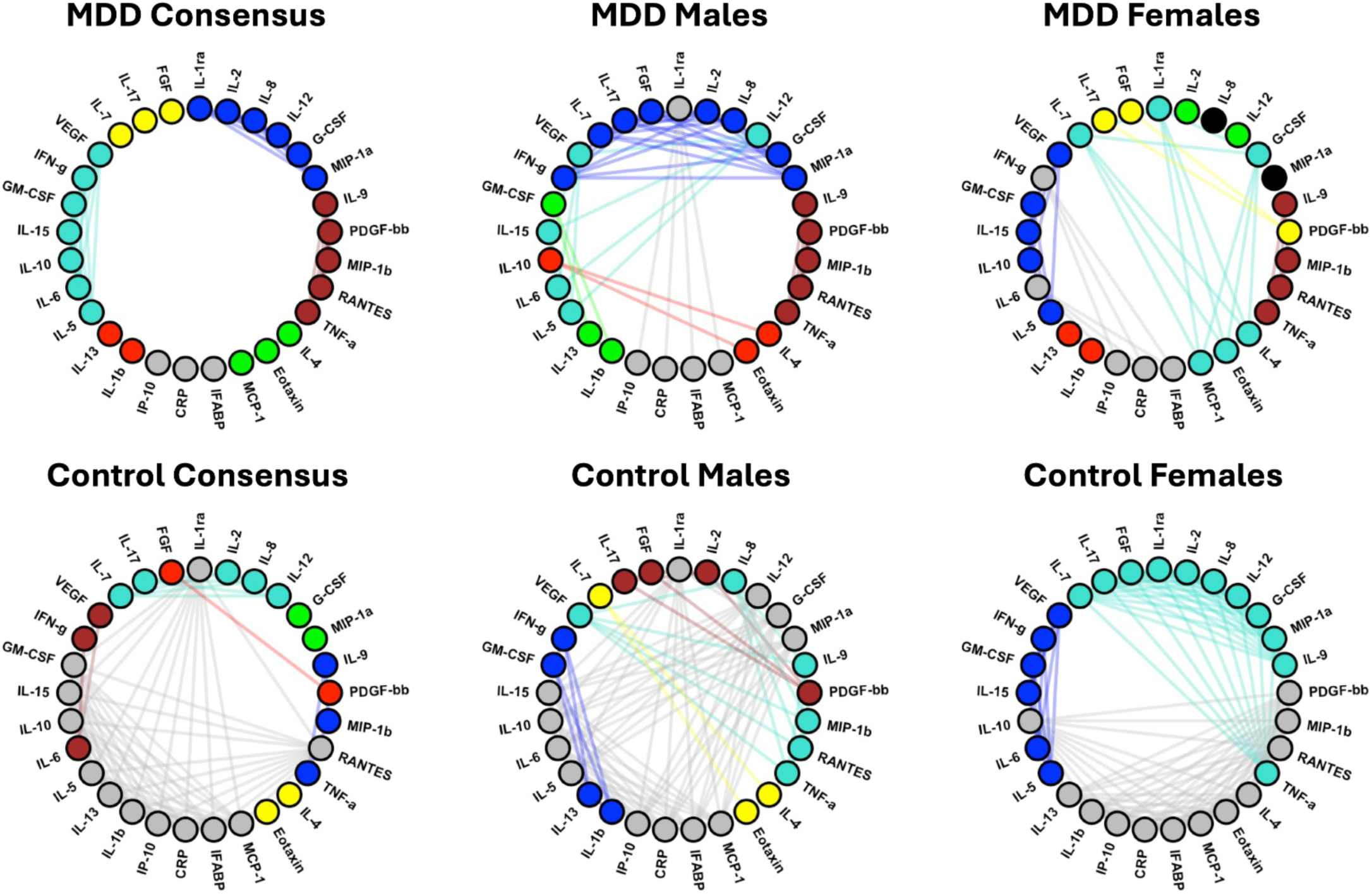
Inflammatory modules for MDD and control groups. Consensus modules consisted of clusters identified for individuals with MDD regardless of sex and healthy controls. Clustering was then applied within sex to identify convergence and divergence of cytokine relationships. In addition to being color coded, the lines connect the cytokines and chemokines that are in the same inflammatory modules. Full lists of cytokines in each module are available in Tables S3-5

We next examined how these same proteins network differently in women or men with or without depression to determine how sex contributed to the formation of networks. In men, several inflammatory modules were moderately conserved between controls and MDD (ARI= 0.229/ NMM= 0.246), including TNF-α/MIP-1β/RANTES- and IL-4/eotaxin-centered modules.

However, these modules were substantially reorganized in MDD. The TNF-α module retained its core composition but exchanged VEGF for PDGF-bb, whereas the FGF/IL-17/IL-2 module expanded through the recruitment of IFN-γ, IL-7, IL-8, G-CSF, and MIP-1α. In addition, several cytokines that were unassigned in controls (grey), including IL-5, IL-6, IL-12, IL-15, IL-10, IL-1β, IL-13, GM-CSF, and IL-1Ra, became incorporated into organized modules in MDD, indicating increased coordination of inflammatory signaling.

In women, a different pattern emerged. Female cytokine networks underwent greater re-organization in the MDD group (ARI = 0.083/ NMM= 0.342). Conserved inflammatory programs included a VEGF-centered module containing GM-CSF, IL-15, and IL-5 and a growth factor module centered on FGF and IL-17. However there was substantial reorganization of cytokine co-expression in the MDD group. TNF-α, MIP-1β, RANTES, and IL-9 formed a distinct inflammatory module, while IL-4, eotaxin, MCP-1, and G-CSF became incorporated into an IL-7-centered module. In contrast, IL-6 and IFN-γ were no longer assigned to a co-expression module, and adaptive immune cytokines that clustered together in controls became partitioned into multiple smaller modules in MDD, indicating sex-specific remodeling of inflammatory network organization.

In addition to demonstrating sex specific cytokine networks, inflammatory network organization differed most substantially between healthy males and females (ARI = -0.038/ NMM= 0.429). Female controls exhibited a large turquoise module containing IL-7, IL-17, FGF, IL-1Ra, IL-2, IL-8, IL-12, G-CSF, MIP-1α, IL-9, and TNF-α, whereas these cytokines were distributed across multiple modules in males. In contrast, males exhibited distinct modules containing FGF, IL-17, IL-2, and PDGF-bb, as well as IL-4, eotaxin, and IL-7, while these cytokines were either incorporated into larger modules or remained unassigned in females. VEGF also demonstrated sex-specific network organization, clustering with TNF-α, RANTES, and MIP-1β in males but with IFN-γ, GM-CSF, IL-15, IL-6, and IL-5 in females. Despite marked differences in cytokine network organization between healthy males and females, several inflammatory programs were conserved across sexes in MDD, including TNF-α/MIP-1β/RANTES/IL-9, FGF/IL-17, VEGF/IL-15/IL-5, and IL-4/eotaxin modules. Consistent with these observations, cytokine network organization was more similar between males and females with MDD than between healthy males and females (ARI= 0.220/ NMM= 0.187), indicating that depression involves a partial convergence of inflammatory networks across the sexes.

After identifying these networks, we examined associations between network-level metrics and group-level MADRS and DARS scores (**Fig. 3**). A positive association indicates that groups with greater network magnitude exhibit higher average scores on the assessment measure, whereas a negative association indicates that greater network magnitude is associated with lower averaged scores. Given that higher DARS scores reflect greater hedonic capacity, positive associations indicate improved hedonic function, whereas negative associations indicate increased anhedonia. For MADRS, where higher scores reflect greater depressive symptom severity, positive associations indicate increased symptom burden. In the MDD consensus group, the brown and yellow modules significantly positively correlated with multiple MADRS symptoms including apparent sadness, reduced sleep and eating along with overall severity.

**Figure 3.**
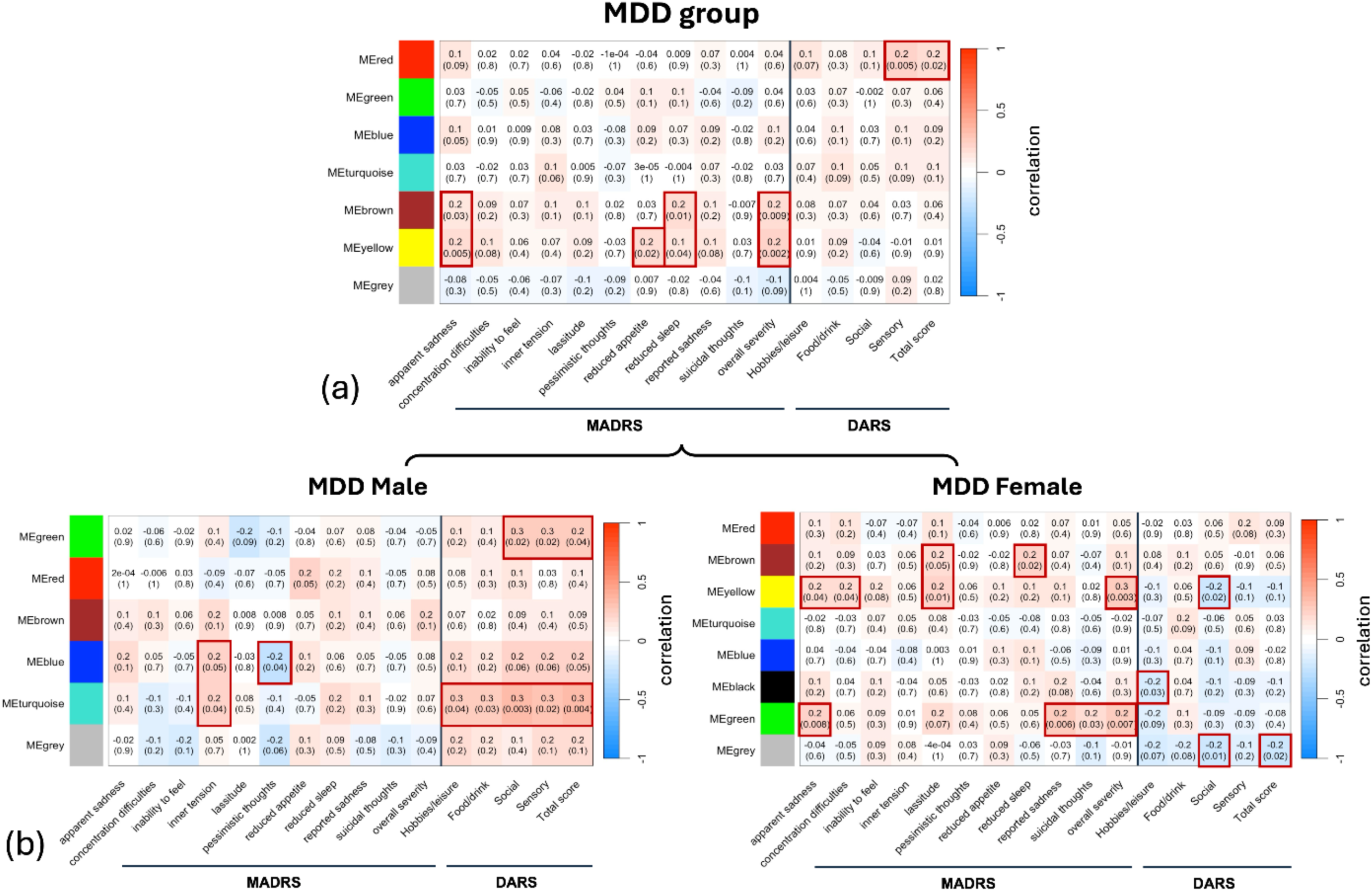
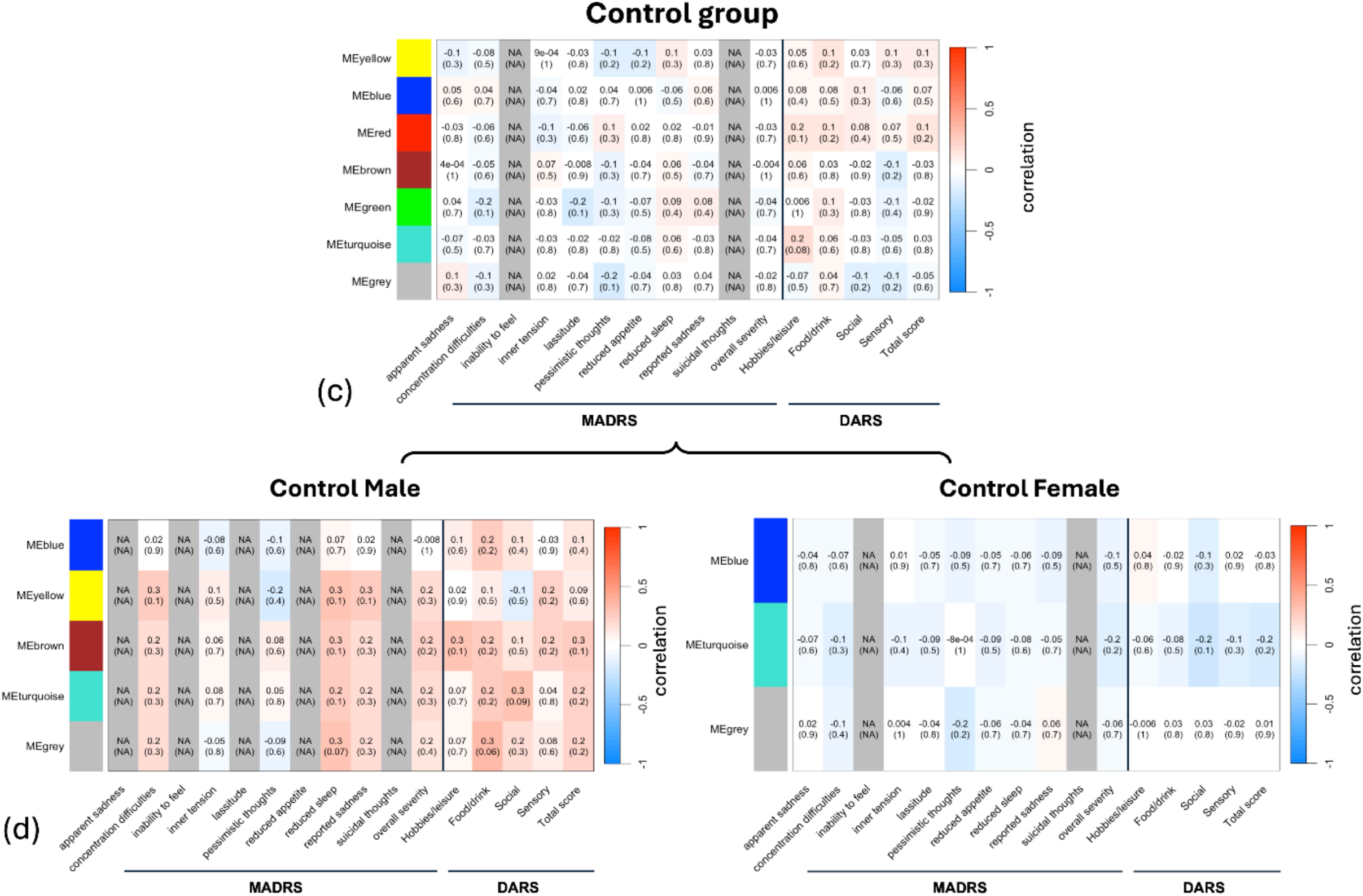
Correlations between blood markers and clinical assessments (MADRS and DARS scores). Inflammatory modules that are male-female consensus are shown in (a) and (c) for the MDD and control groups, respectively. Each group is then subdivided into male and female groups (b and d), and significant correlations with clinical assessments are highlighted in the same manner. In all cases, statistically significant correlations are highlighted by boxes. The numbers outside of the brackets indicate correlation coefficient, and the numbers inside indicate p-value.

We also examined the DARS score to identify which dimensions of anhedonia relate to cytokine networks. When cytokine networks were identified across men and women with MDD we found that the red group made up of IL-13 and IL1β positively correlated to increased sensory and total scores. This positive correlation between sensory and total score to the cytokines IL-13 and IL1β was maintained in men (Green module) but not women. Furthermore when cytokine networks were divided by sex, more male cytokine clusters correlated with DARS categories, whereas clusters of women’s cytokines correlated to a higher number of MADRS categories. The only significant negative correlations between cytokine networks and DARS occurred in women, which reflect that stronger cytokine networks relate to greater anhedonia symptoms (since higher DARS corresponds to greater pleasure).

After identifying inflammatory cytokine modules associated with specific depressive symptoms, we next asked whether these same inflammatory modules were also associated with alterations in brain microstructure measured using diffusion MRI (**Fig. 4**). Among the diffusion MRI measures, CDI and FAt showed the most widespread associations with the inflammatory modules. Higher CDI was associated with elevated eigen-gene values in multiple inflammatory modules across widespread cortical and subcortical regions (**Fig.4**). The strongest associations were observed in frontal and temporal cortical areas, with additional associations in the thalamus, globus pallidus, and anterior corpus callosum. The brown and yellow modules, which were the same inflammatory modules associated with apparent sadness, reduced sleep, and reduced appetite, showed the strongest and most consistent associations with increased CDI in frontal and temporal regions. FAt also showed widespread associations with inflammatory modules, although the regional pattern differed from that observed for CDI. The brown module was associated with lower FAt in the supramarginal and rostral frontal cortices, whereas the yellow module was associated with higher FAt in precentral regions.In contrast to CDI, higher free-water signal fraction (*f*) was associated only with the yellow module, primarily within temporal and occipital regions.

**Figure 4.**
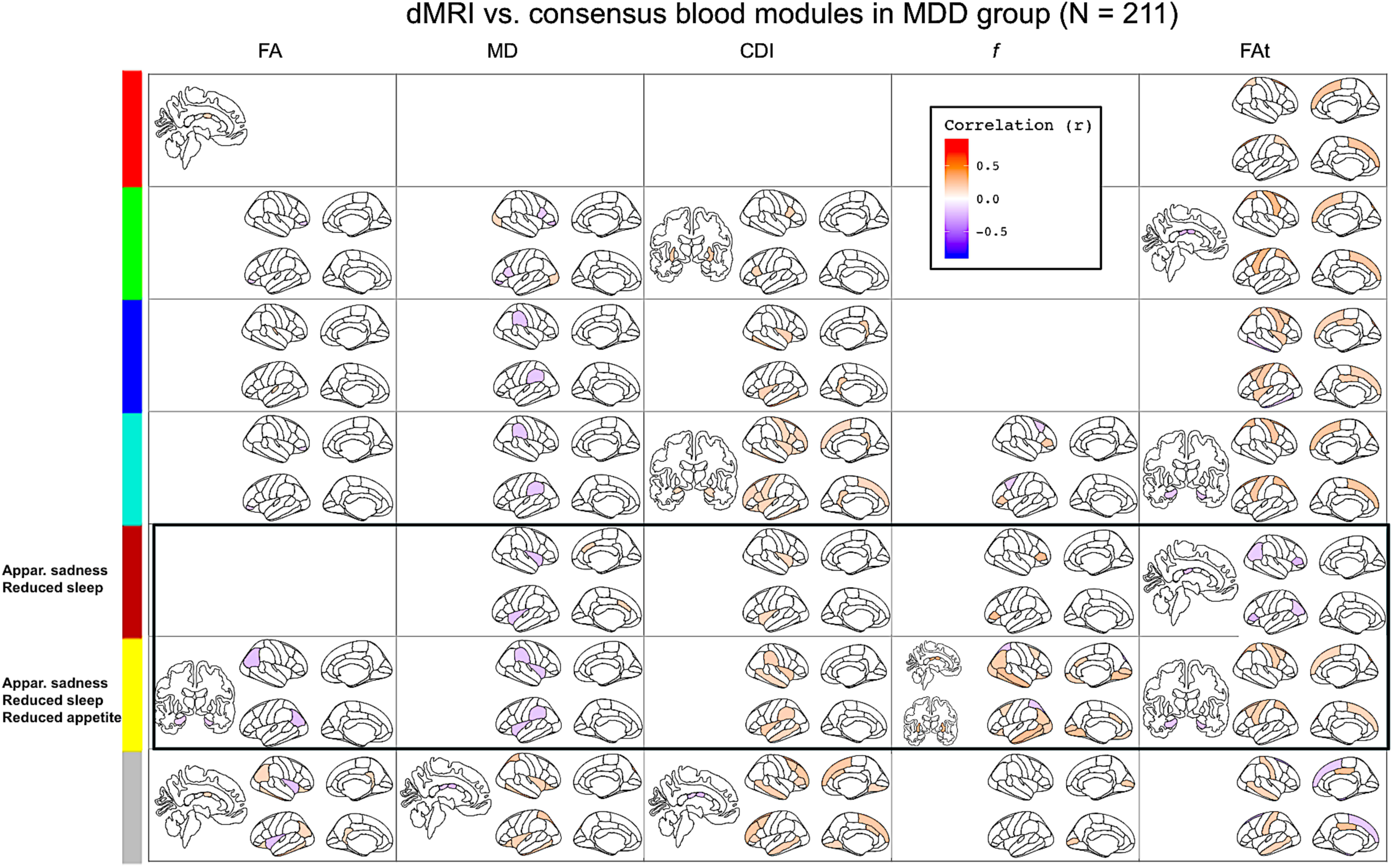
dMRI-blood-module correlation maps for the MDD group. The male-female consensus blood modules are used. The black boxes indicate modules that are significantly correlated with the clinical symptoms listed on the left.

Sex-specific analyses revealed substantially more widespread associations between inflammatory modules and diffusion MRI measures in females than in males **(Fig. 5)**. In females, CDI was predominantly negatively associated with the pro-inflammatory cytokine modules across frontal, temporal, and sensorimotor cortical regions, including the superior temporal gyrus and postcentral gyrus. Many of these same regions also showed negative associations with FA and FAt, whereas MD showed positive associations with inflammation, particularly in the superior frontal and postcentral gyri. The convergence of these diffusion MRI measures within overlapping brain regions suggests that inflammation is associated with widespread alterations in gray matter microstructure in females with MDD.

**Figure 5.**
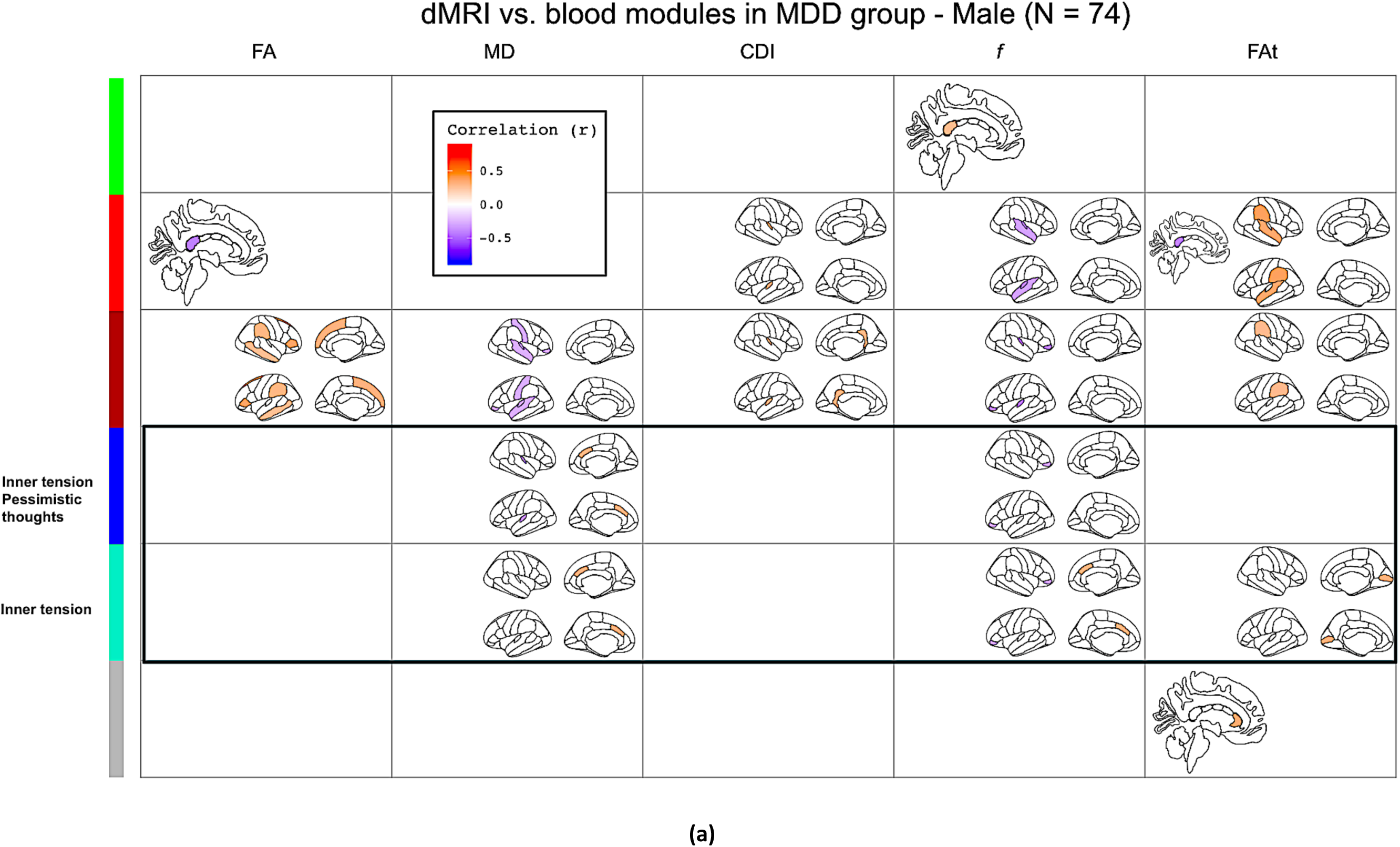
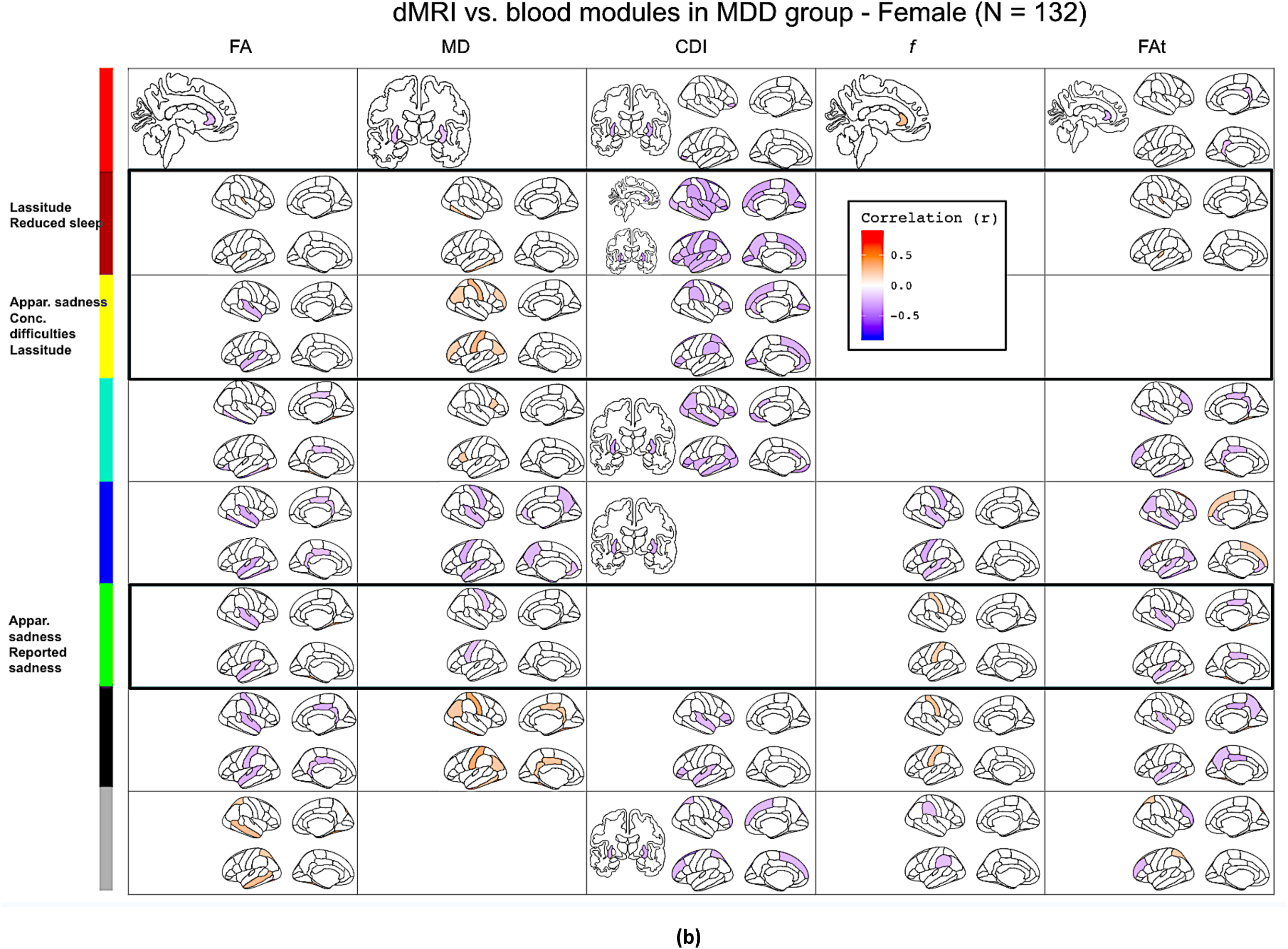
dMRI-blood-module correlation maps for the MDD group, separated by sex. Male control subjects (a) display much fewer dMRI correlations than females (b). The black boxes indicate modules that are significantly correlated with the clinical symptoms listed on the left.

Compared to the grey matter, there were fewer inflammatory-module associations in the white matter. Aside from the anterior corpus callosum (see **Fig. S4**), the only significant association was a positive link between CDI and the turquoise module in the tapetum of the MDD group.

To determine whether these inflammation-related brain changes were unique to depression or reflected more general brain–immune relationships, we compared the regional patterns observed in the MDD and control groups (Table 1). The medial prefrontal cortex, sensory cortex, and temporal cortices were the regions most consistently associated with inflammatory modules in both groups, with additional involvement of the lateral superior frontal cortex. Although similar brain regions were implicated in both MDD and controls, the direction of these associations often differed between groups, suggesting that inflammation may influence brain microstructure differently in health and depression. The widespread CDI associations were consistent with inflammation-related alterations in tissue microstructure, although the specific biological basis of these changes cannot be determined from diffusion MRI alone.

**Table 1.**
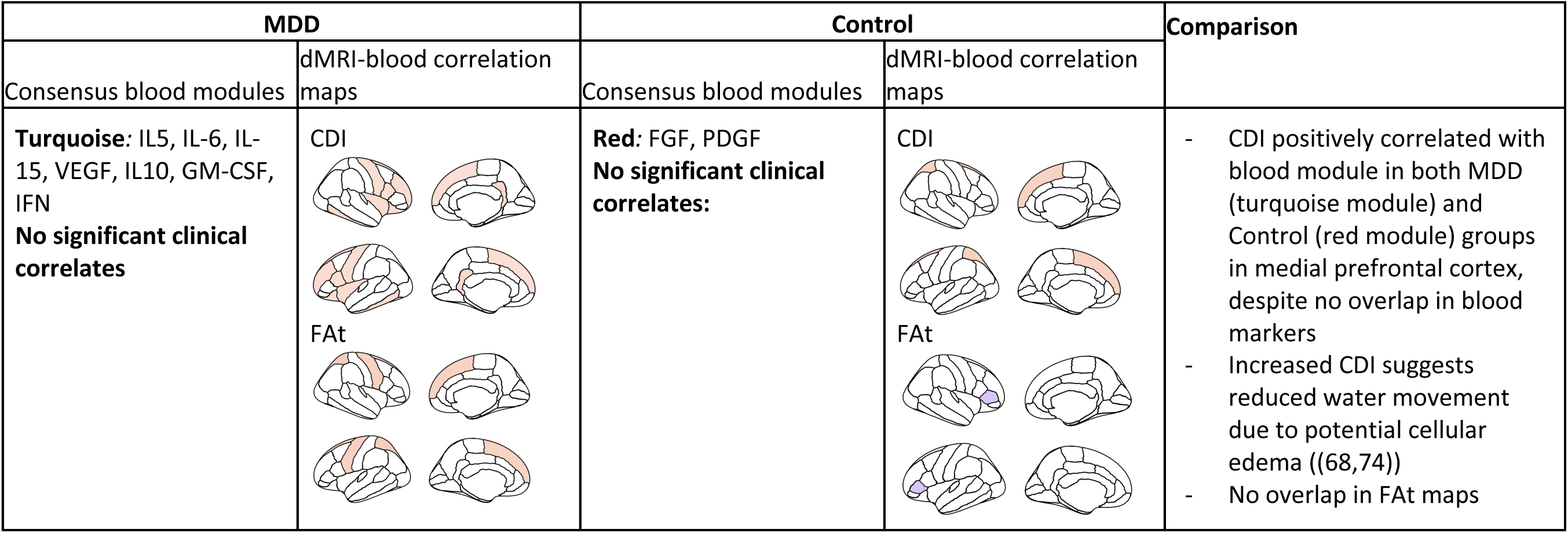
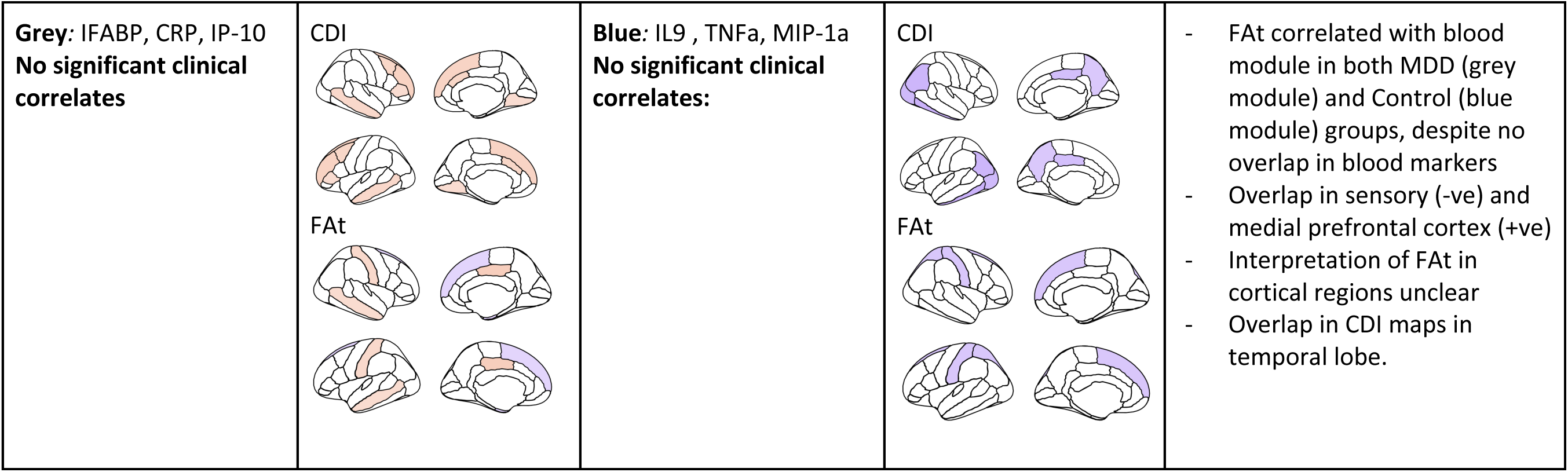
Comparison of inflammatory and clinical associations with imaging markers in common regions: regions that displayed significant correlations in both MDD and control groups.

| MDD |  | Control |  | Comparison |
| --- | --- | --- | --- | --- |
| Consensus blood modules | dMRI-blood correlation maps | Consensus blood modules | dMRI-blood correlation maps |  |
| **Turquoise:** IL5, IL-6, IL-15, VEGF, IL10, GM-CSF, IFN<br>**No significant clinical correlates** | **CDI**<br>**FAt** | **Red:** FGF, PDGF<br>**No significant clinical correlates:** | **CDI**<br>**FAt** | - CDI positively correlated with blood module in both MDD (turquoise module) and Control (red module) groups in medial prefrontal cortex, despite no overlap in blood markers<br>- Increased CDI suggests reduced water movement due to potential cellular edema ((68,74))<br>- No overlap in FAt maps |
| **Grey:** IFABP, CRP, IP-10<br>**No significant clinical correlates** | CDI<br>FAt | **Blue:** IL9 , TNFa, MIP-1a<br>**No significant clinical correlates:** | CDI<br>FAt | - FAt correlated with blood module in both MDD (grey module) and Control (blue module) groups, despite no overlap in blood markers<br>- Overlap in sensory (-ve) and medial prefrontal cortex (+ve)<br>- Interpretation of FAt in cortical regions unclear<br>- Overlap in CDI maps in temporal lobe. |

To determine whether inflammatory cytokine networks link brain microstructure to depressive symptoms, we performed a mediation analysis (**Fig. 6**). In females with MDD, the yellow inflammatory module *(IL-17, FGF, and PDGF)* significantly mediated the relationship between widespread CDI measures across frontal, parietal, temporal, occipital, limbic, and subcortical brain regions and both depression severity (MADRS) and anhedonia (DARS).

**Figure 6.**
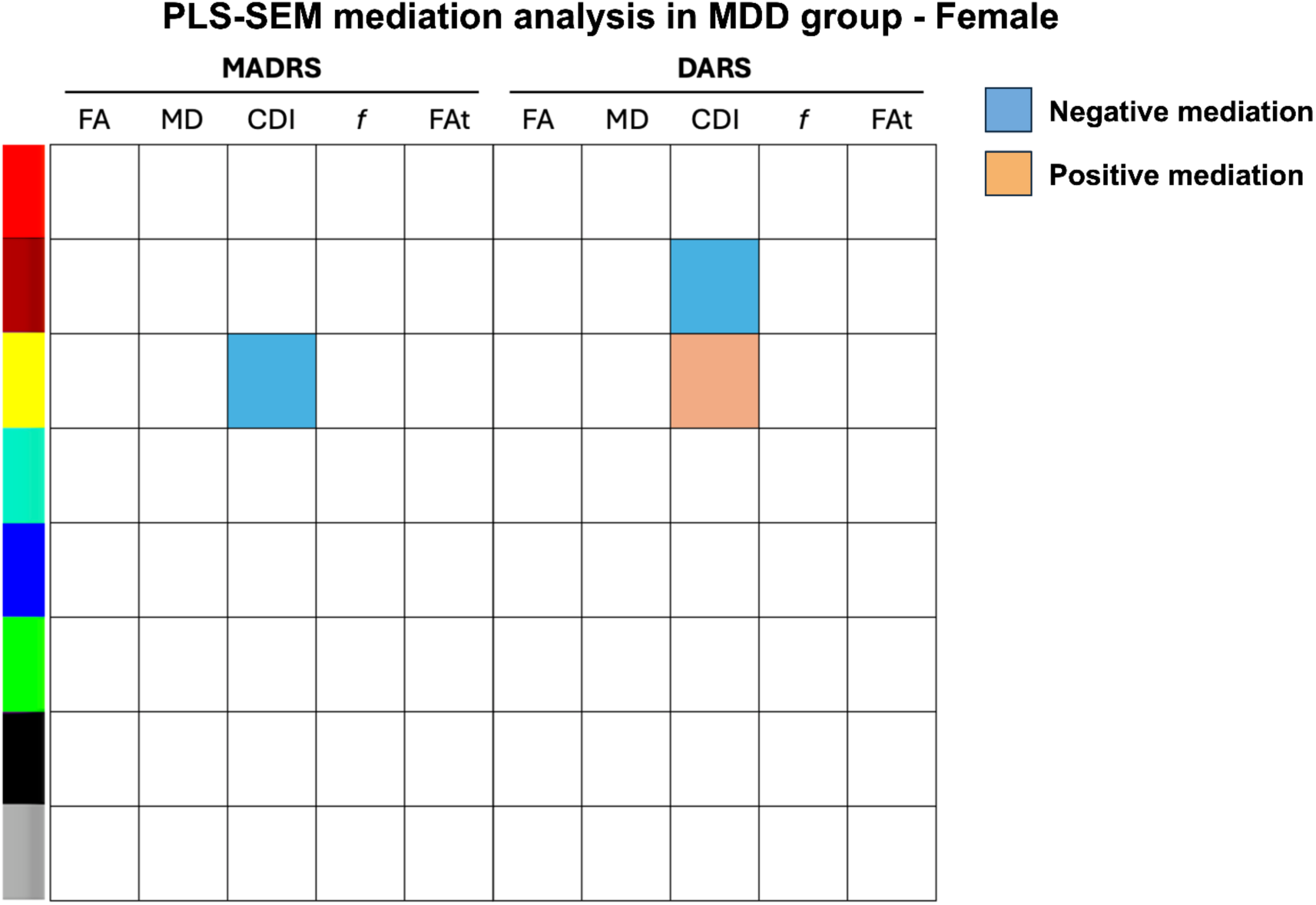
Partial least squares - Structural equation modelling (PLS-SEM) mediation analysis results for the MDD group, Female. Blue (orange) boxes indicate significant negative (positive) indirect effect of the dMRI metrics, mediated through the inflammation modules, on the clinical assessments (MADRS and DARS score).

Specifically, the yellow module mediated a negative association between CDI and MADRS scores (β = -0.14, *p* < 0.05) and a positive association between CDI and DARS scores (β = 0.14, *p* < 0.05). The brown inflammatory module also significantly mediated a negative association between CDI and DARS scores (β = -0.16, *p* < 0.05). No significant mediation effects were observed in males with MDD or in healthy controls.

To determine whether integrating inflammatory, neuroimaging, and clinical measures could identify biologically distinct subtypes of MDD, we performed an agnostic Similarity Network Fusion (SNF) analysis separately in males and females (**Fig. 7**). In males, SNF identified two clusters that primarily differed in the overall magnitude of inflammation, brain microstructural changes, and depression severity. Cluster 1 exhibited higher WGCNA module eigengene values, higher CDI values, and greater MADRS scores than Cluster 2. In females, SNF also identified two clusters; however, unlike males, these groups were not distinguished by MADRS scores. Instead, they differed in WGCNA module eigengene values and regional patterns of brain microstructure. Cluster 1 exhibited higher WGCNA module eigengene values than Cluster 2, whereas CDI and MD showed region-specific increases and decreases rather than a uniform shift across the brain. Together, these findings indicate that integrating inflammatory modules, brain microstructure, and clinical measures identifies distinct subgroups within male and female participants with MDD.

**Figure 7.**
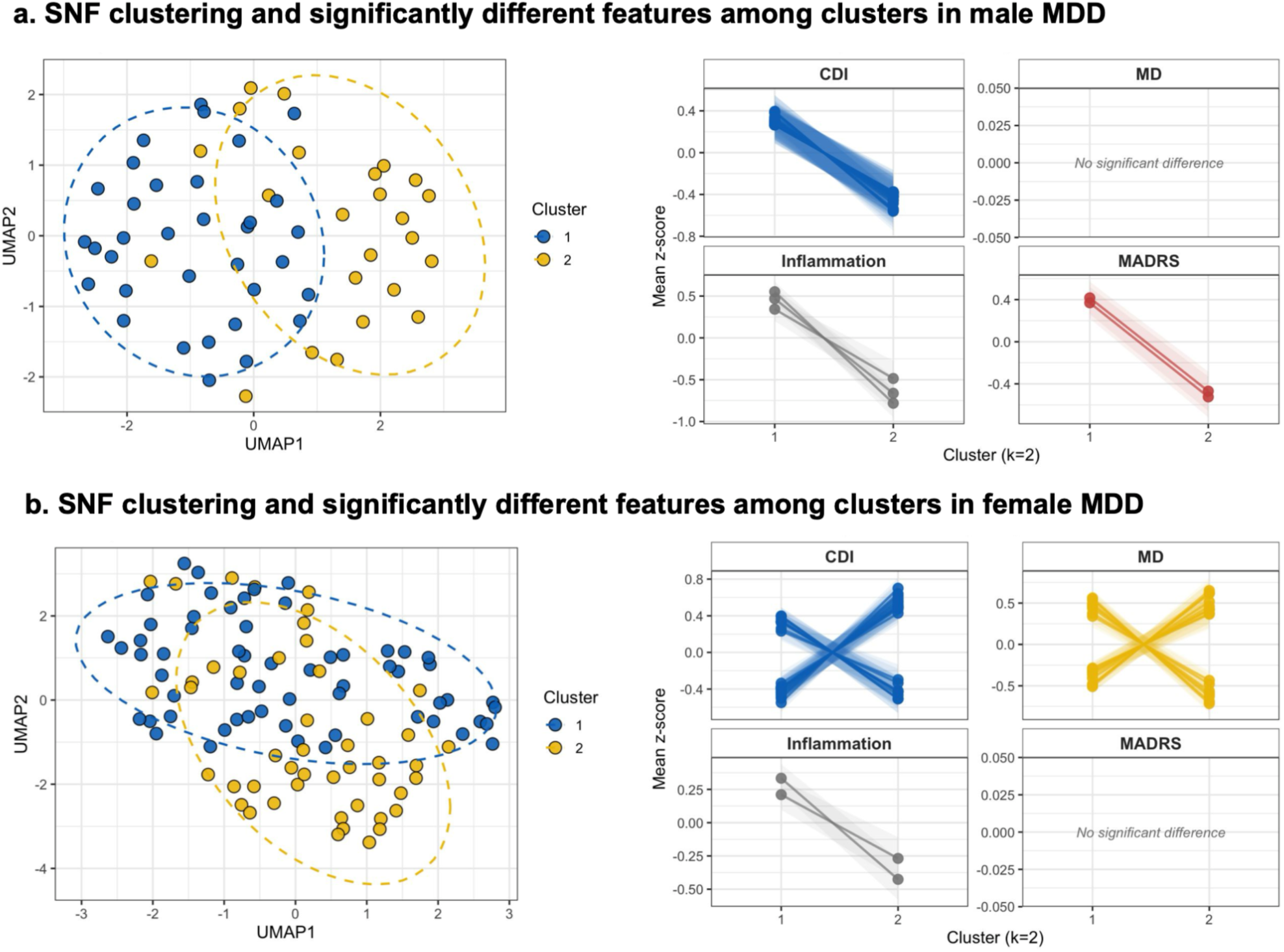
Agnostic clustering of male (a) and female (b) MDD groups was performed using Similarity Network Fusion with multimodal data (CDI, MD, WGCNA inflammation module, and MADRS). UMAP plots (left panel) visualize the two-cluster solution (k=2), with blue representing cluster 1 and yellow representing cluster 2. The right panel shows the mean z-scores for features that differed significantly across clusters, with shading indicating standard errors.

## Discussion

In this exploratory study, we sought to determine whether coordinated networks of peripheral inflammatory cytokines associated with depressive symptoms also correspond to regional differences in brain microstructure, which has been proposed as a means to observe neuroinflammation (19,20,22). Rather than focusing on predefined inflammatory markers, we used an agnostic, data-driven approach to identify cytokine networks based on patterns of coordinated expression across individuals. Our findings support this framework. We found that specific inflammatory cytokine networks were associated with distinct depressive symptoms, widespread alterations in gray matter microstructure, and sex-specific brain–immune relationships. Furthermore, inflammatory cytokine networks statistically mediated the relationship between brain microstructure and both depression severity and anhedonia in females with MDD. Together, these findings support a systems-level model in which coordinated inflammatory networks, rather than individual cytokines, contribute to the biological heterogeneity of depression.

Meta-analyses have consistently demonstrated elevated circulating inflammatory markers in subsets of patients with MDD, supporting a role for immune dysregulation in depression (30–32). While most studies have focused on individual cytokines, more recent agnostic approaches have identified inflammatory subtypes using immune cell populations or aggregate inflammatory measures (33–36). Here, we extend these approaches by identifying coordinated inflammatory cytokine networks and integrating them with symptom dimensions and diffusion MRI measures of brain microstructure. The main findings of this study are: (1) There is a spatially specific association between brain-tissue microstructure and systemic inflammatory markers from the blood, concentrated in the grey matter; (2) these associations are associated with MDD disease severity and are selectively linked to certain symptoms, but do not generalize to the control group; (3) these associations are much more pronounced in female than male MDD patients; (4) of all single-shell dMRI metrics, CDI was most specifically associated with these inflammatory modules; (5) the extent to which symptoms, systemic inflammation and brain microstructure are inter-related demonstrates sex-specific subtypes.

### Inflammatory networks bridge depressive symptoms and brain microstructure

We identified inflammatory cytokine modules (brown and yellow) associated with depressive symptoms in the MDD group, including TNF-α, IL-7, IL-9, FGF, and PDGF as components of these symptom associated modules. No such symptom associated modules were detected in controls, and the composition of the inflammatory networks also differed between MDD and controls.

These data support that both the organization of inflammatory networks and their association with mood-related symptoms are altered in depression. Within the MDD group, inflammatory modules were associated with only a subset of depressive symptoms (**Fig. 3**), particularly apparent sadness, reduced sleep, and reduced appetite, consistent with previous studies linking inflammation to specific symptom dimensions rather than depression as a whole (37–40).

Integrating these inflammatory networks with diffusion MRI demonstrated that the same cytokine modules associated with depressive symptoms also corresponded to widespread alterations in brain microstructure. As shown in **Fig. 4**, CDI showed the most extensive associations with the symptom-specific inflammatory modules, including in the frontal gyrus, the middle temporal gyrus, the frontal pole, the rostral-middle frontal gyrus, the partriangularis, the supramarginal gyrus, the thalamus, the globus pallidus, and the anterior corpus callosum. Notably, these associations were concentrated within gray matter, and the majority of these areas coincide with those reported in the literature (**Fig. 8**), as summarized in **Fig. 9** and **Table 1**) (41–49). Through this imaging integration, our findings provide a potential biological framework for understanding why these regions are vulnerable.

**Figure 8.**
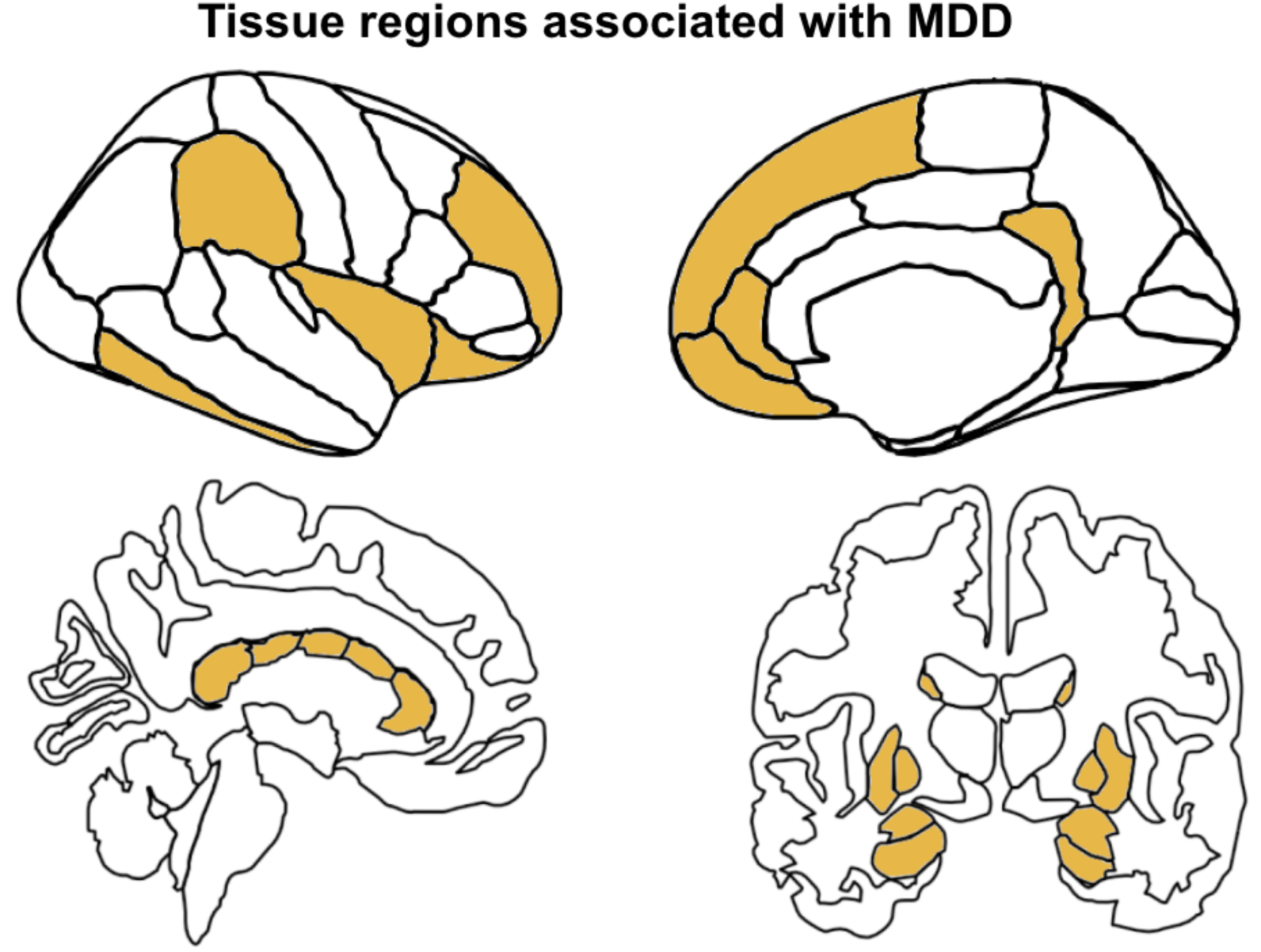
Regions implicated in MDD, based on structural and functional MRI studies and meta-analyses in the literature. These regions, marked in yellow across the two hemispheres as well as in deep brain, include the prefrontofrontal (medial and lateral), anterior cingulate, superior frontal, posterior cingulate, insula and supramarginal regions, as well as the caudate, putamen, hippocampus, amygdala and corpus callosum.

Many of the same cortical regions also exhibited inflammation-related associations in healthy controls, although these relationships were substantially less extensive and were frequently in the opposite direction (**Fig. S4**). Interestingly, in controls CDI was uniquely associated with the blue module (IL-9, TNF-a and MIP-1a), red (FGF, PDGF) and turquoise modules (VEGF, IL-2, IL-7, IL-8, IL-12, IL-17) in the male-female consensus analysis. This difference may highlight the different actions of growth factors compared to dominant microglial activating cytokines such as TNF-a and MIP-1a, and may further highlight the potential sensitivity of CDI to these differences. These findings suggest that immune–brain communication is not only observed in depression but may represent a normal biological process that becomes dysregulated in MDD.

### Sex differences in neuroimmune organization

One of the most striking findings of this study was the consistent evidence for sex differences in neuroimmune organization. Surprisingly, healthy males and females differed most in the organization of peripheral cytokine networks with 42.9% of possible cytokine pair relationships differing between the sexes. In comparison, depression reorganized these networks into more similar inflammatory programs across sexes resulting in only 18.7% of the cytokine pairwise relationships being sex-specific. However, this increased similarity arose through different patterns of network remodeling. Healthy females exhibited relatively large, coordinated cytokine modules that became partitioned into smaller, more specialized co-expression networks in MDD. In contrast, males showed the opposite pattern, with cytokines that were distributed across multiple modules in healthy controls becoming incorporated into larger inflammatory networks in MDD. Several conserved cytokine modules emerged in both male and female MDD, including TNF-α-, VEGF-, FGF-, and IL-4/eotaxin-associated networks, suggesting that depression increased similarity in peripheral cytokine networks without eliminating sex differences in how cytokines were grouped.

Despite this partial convergence of peripheral cytokine networks, brain–immune relationships remained strongly sex dependent. Across multiple independent analyses, inflammatory cytokine networks exhibited stronger associations with brain microstructure in females than in males. Moreover, inflammatory cytokine networks mediated the relationship between brain microstructure and both depression severity and anhedonia exclusively in females. Together, these findings suggest that while depression partially aligns peripheral inflammatory organization across sexes, the relationship between peripheral immune signaling and brain microstructure remains fundamentally different in males and females.

These sex differences were most pronounced in the MDD group. As shown in **Fig. 5**, inflammatory cytokine networks exhibited substantially stronger associations with brain microstructure in females than in males, indicating that the inflammatory–brain relationships observed in the combined MDD cohort (**Fig. 4**) were driven primarily by females. This finding is consistent with the broader literature demonstrating that females are more likely to develop MDD (50–52), experience greater functional impairment (53), and exhibit a more chronic disease course (54). Importantly, the inflammatory networks associated with depressive symptoms also differed between sexes. Rather than implicating the same cytokines, males and females exhibited distinct symptom-associated inflammatory modules that were differentially related to brain microstructure. Together, these findings suggest that sex differences in MDD reflect not only differences in peripheral immune organization but also differences in how peripheral inflammatory networks relate to the brain.

### Grey vs. white matter, across different dMRI metrics

Our study helps to establish feasibility of single-shell dMRI for understanding inflammatory markers in brain microstructure. As briefly alluded to earlier, MD and CDI are often found with contradicting associations with the same blood modules, as a higher MD and a lower CDI are both associated with fast diffusion, and the opposite for slower diffusion. We propose that dMRI markers such as CDI may thus be explored for early detection of inflammation-related brain abnormalities in the context of depressive symptoms. However, we do not presume any of them directly reflects these inflammatory processes, as this goes beyond this proof-of-concept study and requires further validation.

Importantly, we identified consistent associations between peripheral inflammatory cytokine networks and diffusion-derived measures of brain microstructure, with the majority of these relationships occurring in grey matter rather than white matter. Although inflammatory associations within white matter have been reported previously, including in the corpus callosum, our findings suggest that grey matter may be particularly sensitive to peripheral inflammatory signaling in MDD. This observation is consistent with studies of neurodegenerative disorders, where inflammatory processes are frequently associated with alterations in grey matter microstructure (55,56).

Several biological mechanisms could contribute to these grey matter diffusion changes. Neuroinflammatory processes alter the cellular and extracellular environment through changes in glial activation, vascular permeability, and tissue water content, all of which can influence water diffusion (19,20,57). In fact, inflammatory correlates in grey-matter dMRI were suggested as a sign of an intermediate stage between relatively healthy grey matter and lesioned white-matter (58), and may reflect early myelin changes. In this study, as in earlier studies there was an association between peripheral inflammation and dMRI makers in the corpus callosum ((16,59,60). However, the majority of inflammatory diffusion associations were with the grey matter, instead of the white matter. This is consistent with previous findings of inflammatory associations in neurodegenerative diseases (55,56), and is consistent with the notion that grey matter may be particularly susceptible to these processes because of its greater vascular density and cellular complexity. However, diffusion MRI cannot distinguish among these mechanisms, and the observed associations should be interpreted as markers of altered tissue microstructure rather than direct evidence of specific inflammatory processes.

### Mediation effect of systemic inflammation

PLS-SEM identified inflammatory cytokine networks as mediators of the relationship between brain microstructure and depressive symptoms, but only in females with MDD (**Fig. 6**). In particular, the yellow inflammatory module, consisting of IL-17, FGF, and PDGF-bb, mediated the association between CDI measures and depression severity, including symptoms of apparent sadness, reduced sleep, and reduced appetite. These findings suggest that coordinated inflammatory signaling, rather than individual cytokines, may link alterations in brain microstructure to specific symptom dimensions in depression.

The biological composition of this module is consistent with previous evidence implicating these molecules in depression and neurovascular function. IL-17 has been reported to be elevated in MDD, particularly in females (61–63), while FGF and PDGF have been associated with vascular remodeling and blood–brain barrier regulation (64–67). Although these shared biological functions may contribute to their coordinated expression, our findings further suggest that examining these molecules as an integrated network provides greater insight into depression than considering each cytokine independently.

Various DWI metrics have been proposed as in-vivo markers of neuroinflammation. Our analysis supported that CDI demonstrated the strongest and most consistent associations with both inflammatory cytokine networks and clinical symptoms. Although prior studies have suggested that diffusion-derived measures may be sensitive to neurovascular and inflammatory processes, including alterations in blood–brain barrier integrity (68), the biological basis of CDI remains incompletely understood. Consequently, our findings should be interpreted as evidence that CDI is sensitive to inflammation-related alterations in brain microstructure rather than as direct evidence of blood–brain barrier dysfunction. Future studies combining diffusion MRI with complementary measures of vascular integrity will be important for defining the biological processes underlying these associations.

### Agnostic subtyping of male and female MDD using Similarity network fusion

To determine if our systems-level approach of integrating inflammatory cytokine networks, brain microstructure, and clinical symptoms into coordinated neuroimmune relationships could identify biologically distinct subtypes of MDD we used SNF. Rather than classifying patients based on individual cytokines, imaging measures, or symptom scores alone, SNF combined these complementary data to identify patient groups with shared brain–immune profiles.

This approach identified sex-specific neurobiological subtypes. In males, one subtype (Cluster 1) was characterized by concordant elevations in inflammatory cytokine networks, CDI abnormalities, and depressive symptom severity, whereas the second (Cluster 2) exhibited relatively lower values across these measures. These findings suggest that a subset of male patients may represent an inflammation-associated form of depression in which peripheral immune activation, brain microstructural alterations, and clinical symptoms are closely aligned consistent with evidence linking inflammation to MDD severity and microstructure disruption (30,69).

Female subtypes differed in a fundamentally different manner. Rather than separating according to the overall magnitude of inflammation or brain abnormalities, female clusters were distinguished by region-specific patterns of brain microstructure despite similar inflammatory and clinical profiles. This suggests that neuroimmune heterogeneity in females may arise from differences in the spatial organization of brain alterations rather than the overall severity of inflammatory activation. Such patterns could be explained by the region-specific effects of sex hormones on inflammation, brain microstructure, myelination, and microglial function, which introduce greater complexity into female compared to male groups (70–72).

These findings illustrate the value of integrating multiple biological domains to define depression subtypes. Individual cytokines, regional diffusion measures, or symptom scores each capture only one aspect of disease heterogeneity. By combining coordinated inflammatory networks, brain microstructure, and clinical symptoms, SNF identified biologically meaningful patient groups that were not apparent from any single data type alone. Such integrated brain–immune phenotypes may provide a framework for precision psychiatry by identifying patients whose depression is driven by distinct biological mechanisms. This may help explain why anti-inflammatory treatments demonstrate efficacy only in subsets of patients. For example, infliximab, a TNF-α inhibitor, improves depressive symptoms primarily in patients with elevated baseline inflammatory markers (73). Rather than selecting patients using a single biomarker such as CRP, multimodal brain–immune signatures may provide a more comprehensive approach for identifying individuals most likely to benefit from targeted immunomodulatory therapies.

### Limitations

We recognize that the sample sizes in this study are limited, and the diffusion MRI data consists of only a single shell. We took advantage of the availability of blood markers in this arm of CANBIND, and will pursue future analyses with more advanced diffusion models to allow better interpretation of the findings.

#### Conclusions

Inflammation is increasingly recognized as a contributor to depression in a subset of individuals; however, identifying which patients are most likely to benefit from immune-targeted therapies remains a major challenge. Rather than focusing on individual cytokines or neuroimaging measures, our findings demonstrate that integrating inflammatory networks, brain microstructure, and symptom dimensions identifies biologically meaningful subtypes of depression. These multimodal brain–immune signatures differ between the sexes and provide a framework for stratifying patients into biologically defined subgroups that may improve the development and application of precision therapies.

## Data Availability

All data produced in the present study are available upon reasonable request to the authors

## Acknowledgments

This work was supported by the Canadian Institutes of Health Research (CIHR), and the Canada Research Chair (CRC) Program.

