## Supplementary materials for "Mapping brain connections to inflammatory networks in depression identifies sex-specific neuroimmune networks and neurobiological subtypes"

*Co-first authors
✝ Co-senior authors

### Supplementary Methods

#### Weighted gene correlation network analysis

The WGCNA R package provides comprehensive tools for network construction, module detection, and visualization (Langfelder & Horvath, 2008). Briefly, the inflammatory markers were first checked for normality, and the markers with abnormal distributions were normalized. The WGCNA pipeline includes the following steps: (1) construction of the correlation matrix: based on pairwise Pearson's correlations of inflammatory markers, with soft-thresholding to emphasize more robust pairs; (2) network construction: the soft-thresholded correlation matrix is converted to an adjacency matrix, based on which a topological overlap matrix is constructed between adjacency modules (i.e. networks); (3) module detection: markers with high topological overlap are clustered into modules using hierarchical clustering; (4) module eigengenes: the first principal component of each module's expression profiles, summarized as a single value (Desquilles & Musso, 2023). Each group was separated by sex, and WGCNA was performed for each sex. Then, the male-female consensus modules, which are present in both sexes, were isolated by merging the male and female topographical overlap matrices within the MDD and control groups, and emphasizing the common modules across sexes.

#### Imaging markers

Mean diffusivity (MD) and fractional anisotropy (FA) were derived using dtifit in FSL. Free-water corrected FA (FAt) and free-water signal fraction (F) were obtained using free-water elimination (fwDTI), implemented using in-house scripts based on the work of Pasternak et al. (74). Additionally, we computed the correlated diffusion index (CDI) based on our recent work (68,74); a higher CDI reflects restricted diffusion, which could reflect early degeneration or cellular edema ((68,74)). A higher MD and *f* both suggest an increased extent of faster diffusion in, e.g. extracellular space. A higher CDI value indicates reduced diffusivity, thus a potential marker of inflammation. A decreasing FA and FAt are both indicative of reduced fibre integrity, such as driven by myelination.

#### Clinical markers

The primary clinical marker was the Montgomery-Åsberg Depression Rating Scale (MADRS), one of the most widely used and respected clinician-rated tools for quantifying the severity of depressive symptoms. The scale consists of ten items (e.g., apparent sadness, inner tension, reduced sleep, suicidal thoughts), each rated on a 0–6 point scale, yielding a total score from 0 to 60. Crucially, the MADRS is known for its high sensitivity to change, making it the gold standard for monitoring treatment response and symptom progression in clinical trials of antidepressant medications. In addition, we assessed the Dimensional Anhedonia Rating Scale (DARS), which measures anhedonia across reward domains including Hobby, Food, Social Activities and Sensory Experiences. The sleep quality was measured by the Pittsburgh Sleep Quality Index (PSQI), including 7 component aspects (e.g. sleep quality, latency, duration, disturbance). Scores were obtained for both the MDD and control groups. Details of the questionnaire for each score were summarized in **Table S7**.

#### Data integration

Furthermore, we performed a mediation analysis using partial least squares structural equation modelling (PLS-SEM) (Hair et al., 2019) to investigate the indirect effects (mediation) of peripheral inflammatory markers on the relationship between dMRI metrics and clinical symptoms. PLS-SEM analysis was performed using the seminR package in R (Hair et al., 2021) to test the hypothesized pathway shown in **Fig. S4**.

To identify data-driven subtypes within MDD groups, separated by sex, Similarity Network Fusion (SNF) (Wang et al., 2014) was performed using the SNFtool package in R. This approach integrates multimodal data including regional CDI and MD, WGCNA inflammation module, and MADRS. All features were z-score normalized prior to clustering. SNF constructs a similarity network for each modality and iteratively fuses them into a single integrated matrix. The eigen-gap and rotation cost heuristics scores were used to determine optimal cluster number, which showed a two-cluster solution (k=2) for both sexes groups. Spectral clustering was then applied to the fused network to derive cluster labels, and the cluster structure was visualized using Uniform Manifold Approximation and Projection (UMAP). Features that significantly differed between clusters were identified using two-sample t-tests (FDR corrected, $p < 0.05$), and were visualized as mean z-score plots with standard errors.

### Supplementary Results

As shown in **Fig. S4**, the control group also exhibits blood modules that are proinflammatory, as indicated by the purple outlines. CDI is positively associated with the red module in the superior frontal cortex and the anterior corpus callosum, and negatively correlated with the blue module in the inferior parietal, posterior cingulate, precuneus and inferior temporal regions. Like CDI, MD is also positively associated with the red module in the superior frontal cortex, but unlike CDI, MD is negatively associated with the red module. Higher levels of proinflammatory markers are predominantly negatively associated with lower CDI values, consistent with increased extracellular space.

When separated by sex, neither the male nor the female groups display significant dMRI correlations with blood modules. Moreover, while the CDI and MD are strongly positively correlated with the yellow module, which consists of eotaxin and IL7, the MD is negatively associated with the blue module, which consists of the anti-inflammatory IL13 and 3 pro-inflammatory microglial-activation-related markers **Fig. S5a**. It is unclear whether these MD associations are driven by the pro- or anti-inflammatory markers. Nonetheless, CDI is consistently positively associated with the blue and yellow modules in the medial-prefrontal cortex, most consistently so than the other dMRI metrics. The female subjects show strong dMRI-blood correlations in the blue module, which is purely pro-inflammatory. Specifically, MD, not CDI, shows negative correlation with the blue module in the temporal cortex and cingulate cortex.

### Supplementary Materials

**Table S1.** Demographics Tables of MDD and healthy controls.

| Demographic | MDD | Controls | p-value |
|---|---|---|---|
| **Sex** (Male/Female) | 74 / 132 | 38 / 65 | 0.97 |
| **Age** | 35.4 ± 12.7 | 33.2 ± 11.0 | 0.11 |
| **BMI** | 24.4 ± 4.84 | 26.6 ± 6.44 | < 0.001 |
| **Education** | 16.9 ± 2.14 | 18.2 ± 2.27 | < 0.001 |

**Table S2.** Cytokines, target cells, and associated immune function. Cytokines are categorized based on dominant immune pathways. Type 1 immunity corresponds to IFN-γ– and IL-12–driven cellular inflammatory responses, while Type 2 immunity reflects IL-4/IL-5/IL-13–mediated humeral, allergic, and tissue-repair pathways, and Type 3 immunity includes IL-17–driven neutrophilic responses. Regulatory cytokines suppress inflammation, growth factors support vascular or stromal remodeling, and damage markers reflect tissue injury rather than immune signaling.

| Abbrev. | Full Name | Primary Target Cells | Immune function/ Subtype |
|---|---|---|---|
| IL-1β | Interleukin-1 beta | Monocytes, macrophages, neutrophils, CD4+ T cells, endothelial cells, Microglia | Type 1 inflammation/pyrogen |
| IL-13 | Interleukin-13 | Epithelial cells, eosinophils, M2 macrophages | Type 2 immunity |
| IL-2 | Interleukin-2 | T cells (CD4+ CD8+), NK cells, Tregs | Type 1 immunity & type 2 immunity / Regulatory |
| IL-1ra | Interleukin-1 receptor antagonist | Monocytes, endothelial cells, B cells, Microglia | Regulatory |
| IL-8 (CXCL8) | Interleukin-8 | Neutrophils | Type 1 chemokine |
| IL-12 | Interleukin-12 | NK cells, CD 4/ CD8 T+ cells, Th1 cells, Microglia | Type 1 immunity |
| IL-1ra | Interleukin-1 receptor antagonist | Monocytes, endothelial cells, B cells, Microglia | Regulatory |
| GM-CSF | Granulocyte-Macrophage Colony-Stimulating Factor | Monocytes, granulocytes, DC precursors | Type 1 inflammation |
| MIP-1α (CCL3) | Macrophage Inflammatory Protein-1α | Neutrophils, Monocytes, macrophages, NK cells | Type 1 chemokine |
| IL-4 | Interleukin-4 | B cells, Th2 cells, eosinophils, M2 macrophages, Microglia | Type 2 immunity |

| | | | |
|---|---|---|---|
| Eotaxin (CCL11) | Eosinophil chemotactic protein | Eosinophils, Basophils, Mast cells | Type 2 chemokine |
| MCP-1 (CCL2) | Monocyte Chemoattractant Protein-1 | Monocytes | Type 1 chemokine |
| IL-5 | Interleukin-5 | Eosinophils, Basophils, B cells | Type 2 immunity |
| IL-6 | Interleukin-6 | Hepatocytes, monocytes, T cells, B cells, Microglia | Pleiotropic-Type 1 |
| IFN-γ | Interferon-gamma | Macrophages, NK cells, T cells, Microglia | Type 1 immunity |
| G-CSF | Granulocyte Colony-Stimulating Factor | Neutrophils, monocytes | Type 1 inflammation |
| IL-10 | Interleukin-10 | Macrophages, monocytes, dendritic cells, T cells, Microglia | Regulatory |
| IL-15 | Interleukin-15 | NK cells, CD8 T memory cells | Pleiotropic-Type 1 |
| VEGF | Vascular Endothelial Growth Factor | Endothelial cells | Growth factor |
| IL-17A | Interleukin-17 | Epithelial cells, fibroblasts, Naïve CD4 T+ cells | Type 3 immunity |
| FGF | Fibroblast Growth Factor | Fibroblasts, endothelial cells | Growth factor |
| IL-9 | Interleukin-9 | Mast cells, T cells, B cells, ILC2 | Type 2 allergic/mucosal |

| | | | |
|---|---|---|---|
| PDGF | Platelet-Derived Growth Factor | Fibroblasts, smooth muscle cells | Growth factor |
| TNF-α | Tumor Necrosis Factor-α | Endothelial cells, monocytes, macrophages, neutrophils | Type 1 inflammation |
| MIP-1β (CCL4) | Macrophage Inflammatory Protein-1β | Monocytes, macrophages, T cells | Type 1 chemokine |
| RANTES (CCL5) | Regulated upon Activation, Normal T cell Expressed and Secreted | T cells, eosinophils, basophils, NK cells, Microglia | Type 1 chemokine |

Table S3 Cytokines clustered by WGCNA in depressed people and healthy controls (men and women combined).

| Module Color | MDD Consensus | Control Consensus |
|---|---|---|
| **Turquoise** | VEGF, IFNγ, GM-CSF, IL-15, IL-10, IL-6, IL-5 | IL-7, IL-17, IL-2, IL-8, IL-12 |
| **Blue** | IL-1Ra, IL-2, IL-8, IL-12, G-CSF, MIP-1α | IL-9, MIP-1β, TNF-α |
| **Brown** | IL-9, PDGF-bb, MIP-1β, RANTES, TNF-α | IL-6, IFNγ, VEGF |
| **Yellow** | FGF, IL-17, IL-7 | IL-4, Eotaxin |
| **Green** | IL-4, Eotaxin, MCP-1 | G-CSF, MIP-1α |
| **Red** | IL-13, IL-1β | FGF, PDGF-bb |
| **Grey** | IFABP, CRP, IP-10 | MCP-1, IFABP, CRP, IP-10, IL-1β, IL-13, IL-5, IL-10, IL-15, IL-1Ra, RANTES, GM-CSF |

Table S4 Cytokine clusters in men with depression and healthy controls

| Module Color | Male MDD | Male Controls |
|---|---|---|
| **Turquoise** | IL-5, IL-6, IL-12, VEGF, IL-15 | IL-8, IL-9, MIP-1β, RANTES, TNF-α, VEGF |
| **Blue** | IFN-γ, IL-7, IL-17, FGF, IL-2, IL-8, G-CSF, MIP-1α | IFN-γ, GM-CSF, IL-13, IL-1β |
| **Brown** | IL-9, PDGF-bb, MIP-1β, RANTES, TNF-α | FGF, IL-17, IL-2, PDGF-bb |
| **Green** | IL-1β, IL-13, GM-CSF, IL-1Ra | — |
| **Yellow** | — | IL-4, Eotaxin, IL-7 |
| **Red** | IL-4, Eotaxin, IL-10 | — |
| **Grey** | MCP-1, IFABP, CRP, IP-10 | IL-1Ra, IL-12, G-CSF, MIP-1α, MCP-1, IFABP, CRP, IP-10, IL-5, IL-10, IL-6, IL-15 |

Table S5 Cytokine clusters in women with depression and healthy controls

| Module color | Female MDD | Female controls |
|---|---|---|
| **Turquoise** | IL-1Ra, G-CSF, IL-4, Eotaxin, MCP-1, IL-7 | IL-7, IL-17, FGF, IL-1Ra, IL-2, IL-8, IL-12, G-CSF, MIP-1α, IL-9, TNF-α |

| | | |
|---|---|---|
| **Blue** | VEGF, GM-CSF, IL-15, IL-10, IL-5 | VEGF, IFN-γ, GM-CSF, IL-15, IL-6, IL-5 |
| **Brown** | IL-9, MIP-1β, RANTES, TNF-α | — |
| **Green** | IL-2, IL-12 | — |
| **Yellow** | IL-17, FGF, PDGF-bb | — |
| **Red** | IL-13, IL-1β | — |
| **Black** | IL-8, MIP-1α | — |
| **Grey** | IFABP, CRP, IP-10, IL-6, IFN-γ | PDGF-bb, MIP-1β, RANTES, IL-4, Eotaxin, MCP-1, IFABP, CRP, IP-10, IL-1β, IL-13, IL-10 |

Table S6 Adjusted RAND Index was used as a measure of similarity of cytokine clusters. Normalized Mirkin metric was used as a measure of differences of cytokine clusters

| Comparison | Adjusted Rand Index (ARI) | Pair Disagreements (of 406) | Normalized Mirkin Metric (NMM) |
|---|---|---|---|
| **Consensus Control vs. Consensus MDD** | 0.175 | 97 | 0.239 |
| **Male Control vs. Male MDD** | 0.2292 | 100 | 0.246 |
| **Female Control vs. Female MDD** | 0.083 | 139 | 0.342 |
| **Male Control vs. Female Control** | -0.038 | 174 | 0.429 |
| **Male MDD vs. Female MDD** | 0.220 | 75 | 0.187 |

**Table S7.** Details of questionnaires used in clinical scores assessment

| Score | Item Name | Question text |
|---|---|---|
| MADRS | Apparent Sadness | Representing despondency, gloom and despair. (More than just ordinary transient low spirits) reflected in speech, facial expressions, and posture. |
| | Concentration Difficulties | Representing difficulties in collecting one's thoughts mounting to incapacitating lack of concentration. |
| | Inability to feel | Representing the subjective experience of reduced interest in the surroundings, or activities that normally give pleasure. The ability to react with adequate emotion to circumstances or people is reduced. |
| | Inner Tension | Representing feelings of ill-defined discomfort, edginess, inner turmoil, mental tension mounting to either panic, dread, or anguish. |
| | Lassitude | Representing a difficulty getting started, or slowness initiating and performing everyday activities. |
| | Pessimistic Thoughts | Representing thoughts of guilt, inferiority, self-reproach, sinfulness, remorse, and ruin. |
| | Reduced Appetite | Representing the feeling of a loss of appetite compared with when well. |
| | Reduced Sleep | Representing the experience of reduced duration or depth of sleep compared to the subject's own normal pattern when well. |
| | Reported Sadness | Representing reports of depressed mood, regardless of whether it is reflected in appearance or not. Includes low spirits, despondency or the feeling of being beyond help and without hope. |
| | Suicidal Thoughts | Representing the feeling that life is not worth living, that a natural death would be welcome, suicidal thoughts, and preparation for suicide. Suicidal attempts should not in themselves influence the rating. |
| | Overall severity | Total Score of all 10 items |
| DARS | DARS_B_1 | 1. I would enjoy these activities |
| | DARS_B_2 | 2. I would have a desire to participate in these activities |
| | DARS_B_3 | 3. I would spend time doing these activities |
| | DARS_B_4 | 4. I want to do these activities |
| | DARS_B_5 | 5. These activities would interest me |
| | DARS_B_6 | 6. These activities would give me pleasure |
| | DARS_B_7 | 7. I would start these activities without being pushed or encouraged |
| | DARS_B_8 | 8. I would begin doing them on my own |

| | | |
|---|---|---|
| | DARS_B_9 | 9. I would do them until it was time to stop |
| | DARS_D_10 | 10. I would make an effort to get/make these foods/drinks |
| | DARS_D_11 | 11. I would enjoy these foods/drinks |
| | DARS_D_12 | 12. I want to have these foods/drinks |
| | DARS_D_13 | 13. I would eat as much of these foods as I could |
| | DARS_D_14 | 14. I would make an effort to eat/drink these foods/drinks |
| | DARS_D_15 | 15. I would actively try to get these foods/drinks |
| | DARS_F_16 | 16. Spending time doing these things would make me happy |
| | DARS_F_17 | 17. I would be interested in doing things that involve other people |
| | DARS_F_18 | 18. I would be the one to plan these activities |
| | DARS_F_19 | 19. I would feel cheerful from participating in these social activities |
| | DARS_F_20 | 20. I would actively participate in these social activities |
| | DARS_F_21 | 21. I would try to seek out these activities |
| | DARS_H_22 | 22. I would actively seek out these experiences |
| | DARS_H_23 | 23. I get excited thinking about these experiences |
| | DARS_H_24 | 24. If I were to have these experiences I would savor every moment |
| | DARS_H_25 | 25. I want to have these experiences |
| | DARS_H_26 | 26. I would make an effort to spend time having these experiences |
| | DARS_Hobbies_Tot | Hobbies/leisure domain subscale (DARS_B_1 + DARS_B_3 + DARS_B_4 + DARS_B_5) |
| | DARS_Food_Tot | Food/drink domain subscale (DARS_D_10 + DARS_D_11 + DARS_D_12 + DARS_D_13) |
| | DARS_Social_Tot | Social domain subscale (DARS_F_16 + DARS_F_17 + DARS_F_18 + DARS_F_20) |
| | DARS_Sense_Tot | Sensory experience subscale (DARS_H_22 + DARS_H_23 + DARS_H_24 + DARS_H_25 + DARS_H_26) |
| | DARS_Tot | Total Score |

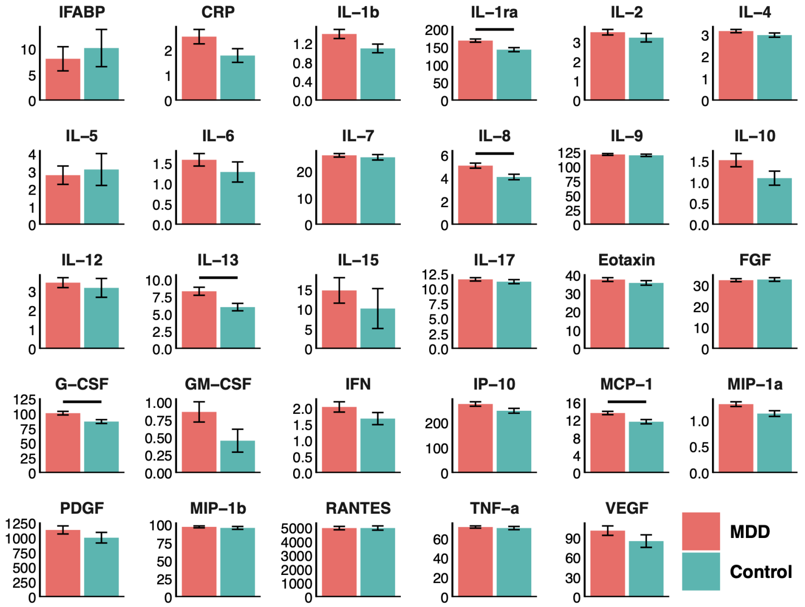
IFABP
CRP
IL–1b
IL–1ra
IL–2
IL–4
IL–5
IL–6
IL–7
IL–8
IL–9
IL–10
IL–12
IL–13
IL–15
IL–17
Eotaxin
FGF
G–CSF
GM–CSF
IFN
IP–10
MCP–1
MIP–1a
PDGF
MIP–1b
RANTES
TNF–a
VEGF
MDD
Control

**(a)**

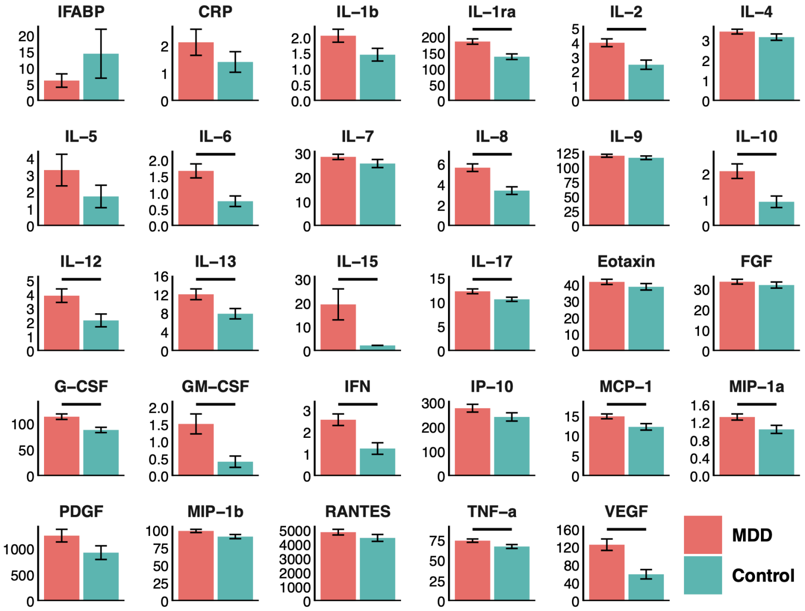
IFABP
CRP
IL–1b
IL–1ra
IL–2
IL–4
IL–5
IL–6
IL–7
IL–8
IL–9
IL–10
IL–12
IL–13
IL–15
IL–17
Eotaxin
FGF
G–CSF
GM–CSF
IFN
IP–10
MCP–1
MIP–1a
PDGF
MIP–1b
RANTES
TNF–a
VEGF
MDD
Control

**(b)**

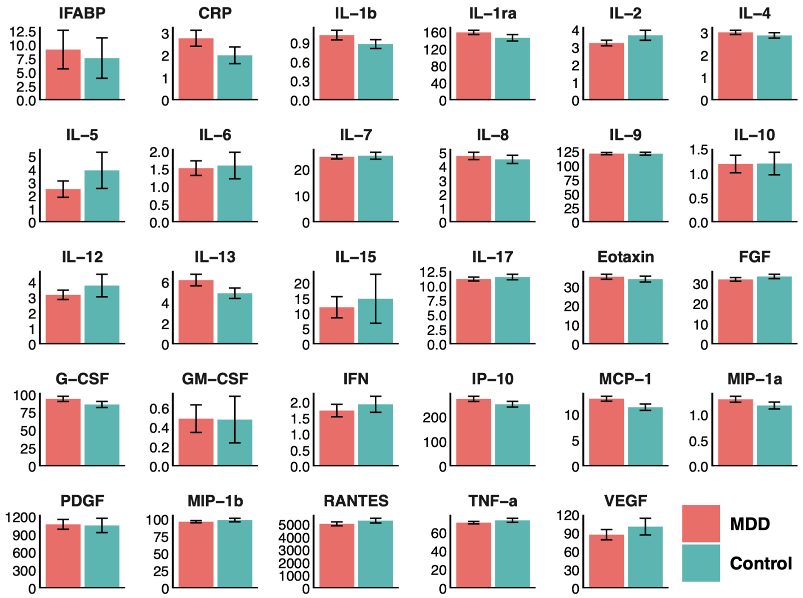


**(c)**

**Figure S1. MDD-Control differences in molecular markers.** The means of all blood makers are plotted for the MDD and control groups, shown in red and green, respectively. (a) is the plot that includes all subjects, (b) and (c) are the plots split by sex, male and female, respectively. Solid horizontal bars represent FDR-adjusted significant differences.

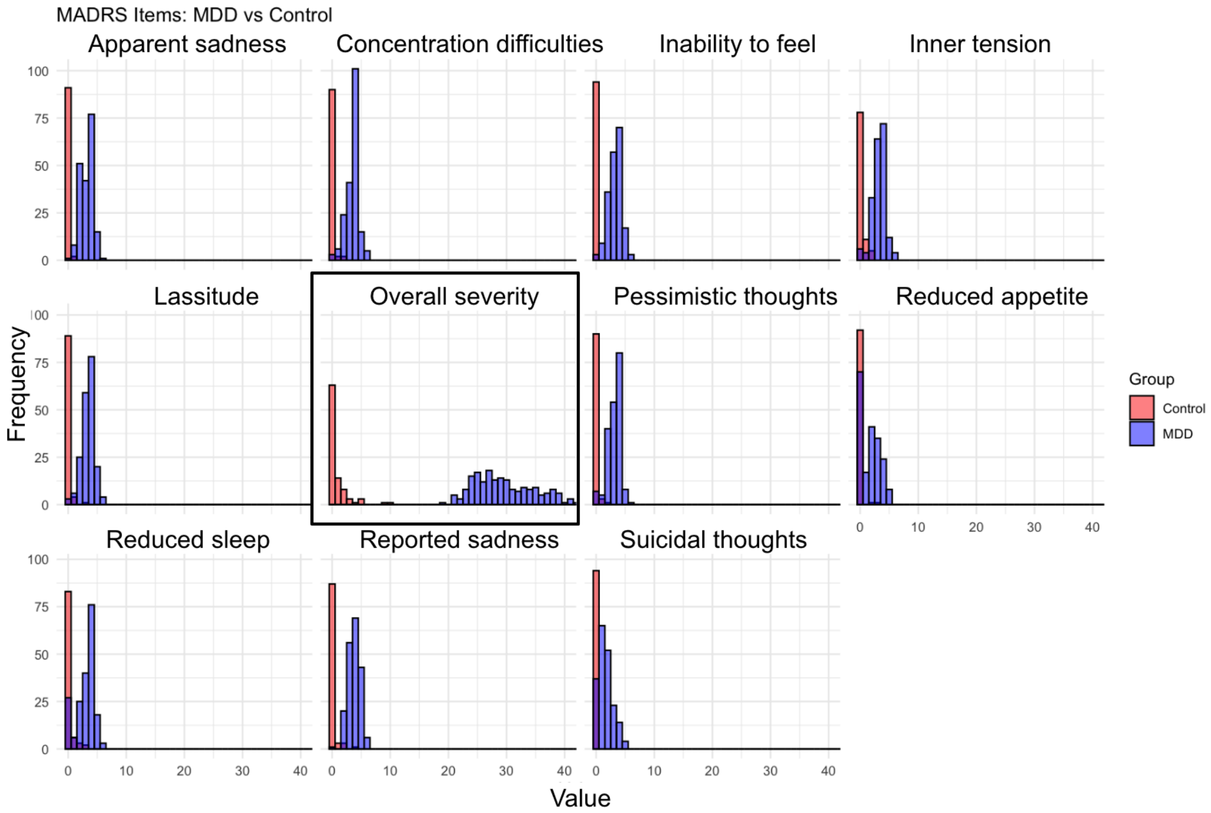


**Figure S2. Distributions of MADRS scores across MDD and Control groups.**

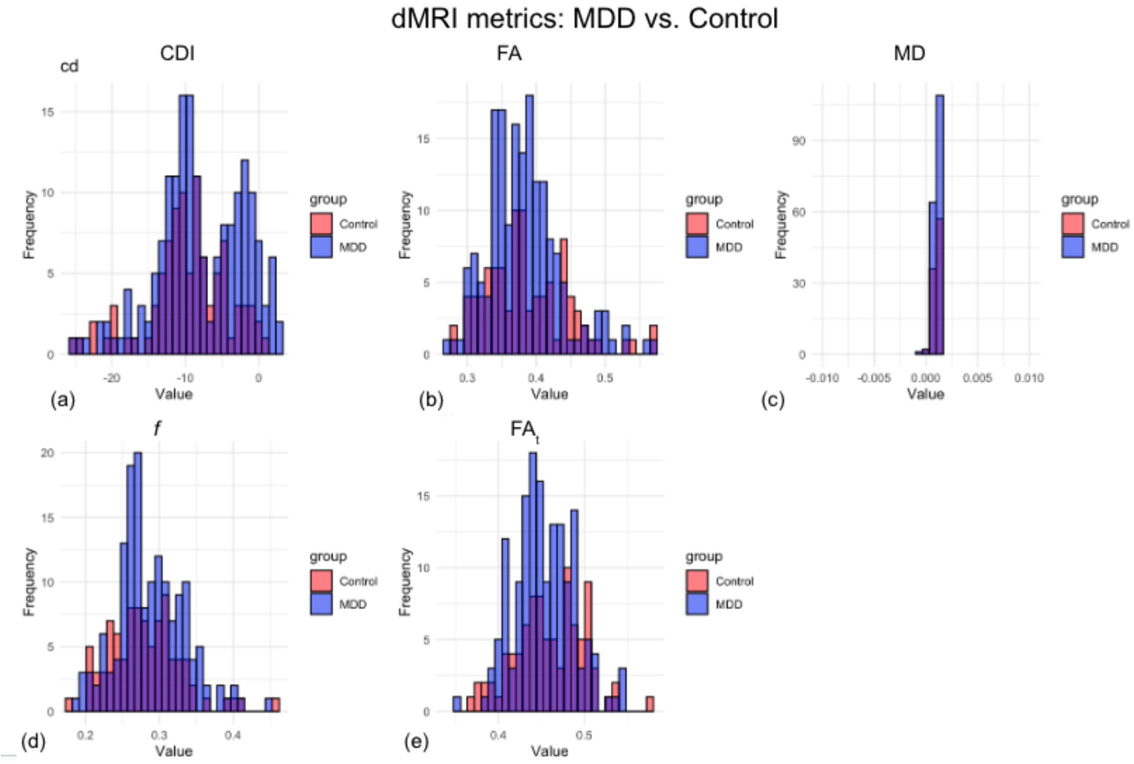


**Figure S3. Distributions of dMRI metrics across MDD and Control groups.**

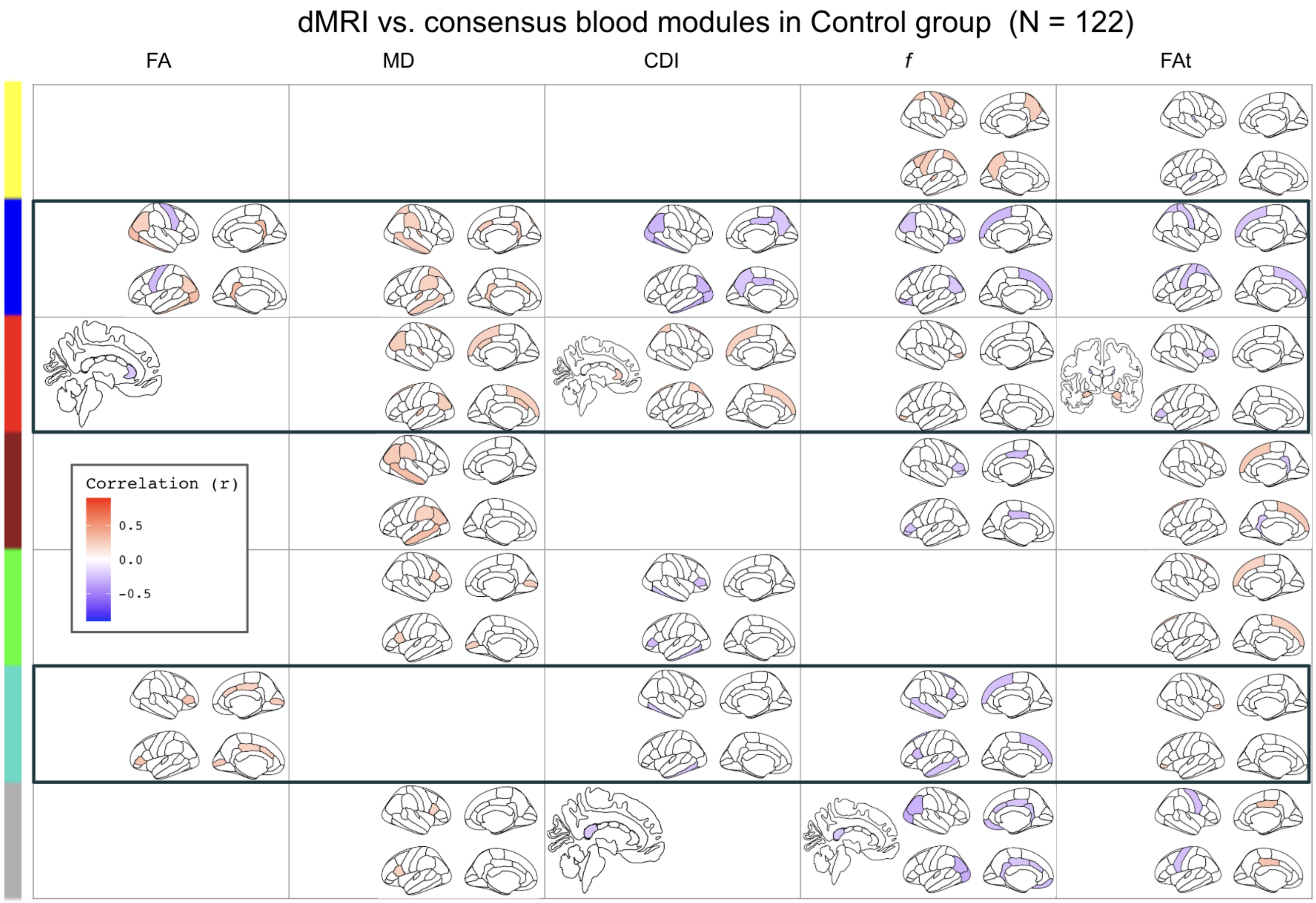


**Figure S4. dMRI-blood-module correlation maps for the control group.** The male-female consensus blood modules are used. The black boxes indicate modules that are significantly correlated with the clinical symptoms listed on the left.

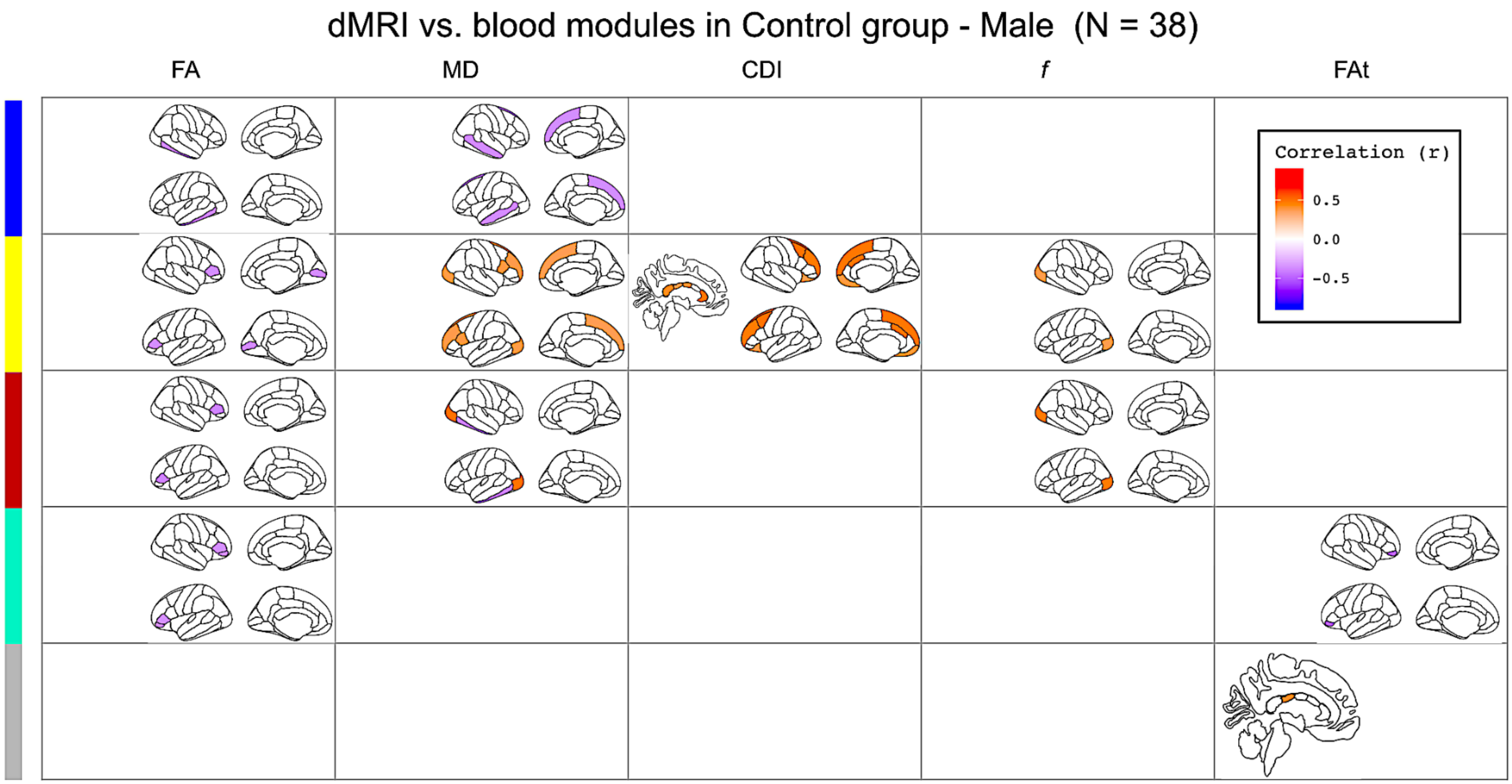
dMRI vs. blood modules in Control group - Male (N = 38)
FA
MD
CDI
f
FAt
Correlation (r)
0.5
0.0
-0.5

**(a)**

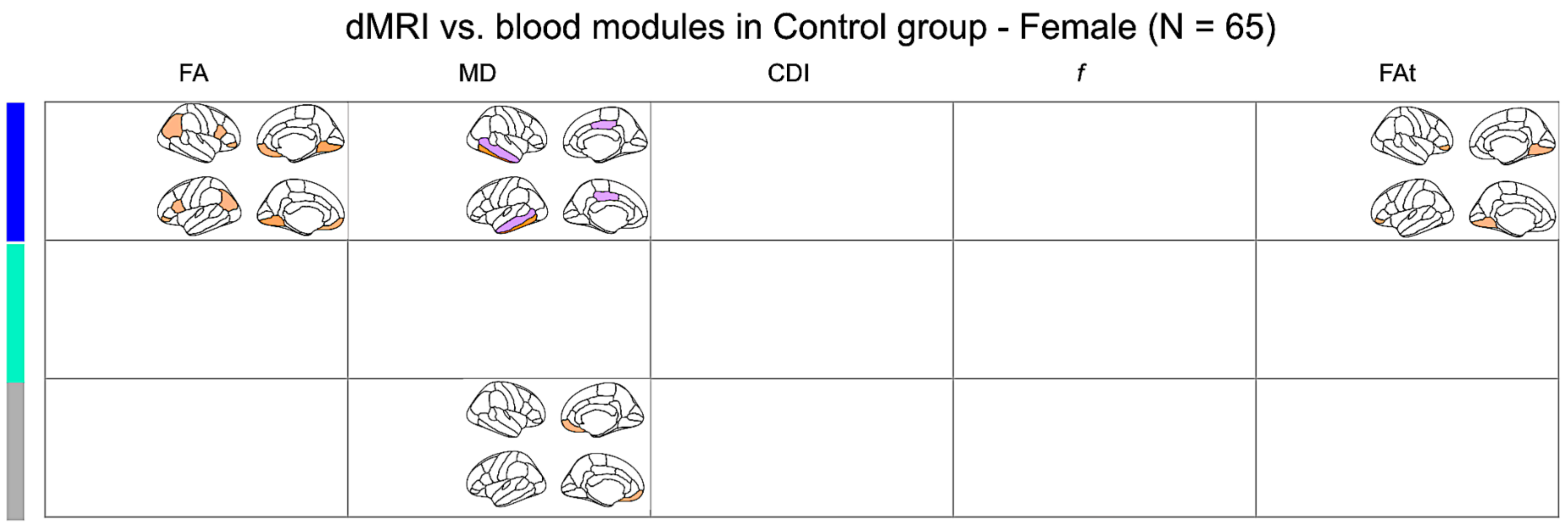


**(b)**

**Figure S5. dMRI-blood-module correlation maps for the Control group, separated by sex.** Male control subjects (a) display more dMRI correlations than females (b).
